# AI-powered single-cell imaging and morphology-guided label-free enrichment reveal tumor heterogeneity, genomic relatedness, and divergence in malignant effusions

**DOI:** 10.64898/2026.11.03.26367106

**Authors:** Mark Jesus M. Magbanua, Laura A. Huppert, Janifer Cruz, Jackson Goudreau, Shaopu Zhou, Keyi Yin, Pavel Stejskal, Natalie Gill, Ryan Carelli, Julie Kim, Linda Hsie, Maggie Wang, Zhouyang Lian, Suchitra Manepalli, Michael Phelan, Manisha Ray, Chassidy Johnson, Jordan Nieto, Emilie Claire Schneider, Anastasia Mavropoulos, Amy L. Delson, Carol Simmons, Amirabbas Parizadeh, Jayla Crawford, Michael Bruck, Lindy Her, Victoria Barnett, Ana Marielle Cabrera, Michelle Melisko, Ronald Balassanian, Hope S. Rugo, Laura van ’t Veer, Andreja Jovic, W. Patrick Devine, Rosalyn W. Sayaman, Mahyar Salek, Maddison Masaeli, Jennifer Rosenbluth, Hani Goodarzi

## Abstract

**Background:** Malignant effusions are clinically accessible yet underutilized sources of metastatic tumor cells. However, abundant background immune cells complicate tumor cell isolation and molecular characterization. We integrate AI-powered single-cell imaging and morphology-guided label-free tumor cell enrichment with genomic profiling to characterize phenotypic and genomic heterogeneity in malignant effusions.

**Methods:** We applied a pre-trained deep learning model to brightfield images of single cells from malignant effusions of 10 patients with metastatic breast cancer to extract morphological features, quantify tumor cell fraction (TCF), and guide label-free microfluidic enrichment. We profiled copy-number alterations and somatic mutations in enriched tumor cell fractions and compared them with available matched effusion cell-free DNA and solid tumors.

**Results:** AI-powered single-cell morphological analysis identified putative tumor cells in all 10 patients (median TCF, 5.4%; range, 0.51–15.8%; n = 25 samples). AI estimates of major immune-cell populations varied across patients and samples and correlated with flow cytometry measurements (n = 21 paired samples, Pearson r = 0.69–0.75). Unsupervised analysis identified four tumor cell morphotypes, revealing intra- and inter-patient morphological heterogeneity and longitudinal variation in morphotype composition. Genomic profiling of enriched tumor cell fractions by low-pass whole-genome sequencing (n = 26 samples) and whole-exome sequencing (n = 24 samples) identified copy-number alterations and somatic mutations, with tumor fraction influencing genomic signal strength. Comparisons of genomic profiles from tumor cell-enriched fractions with matched effusion cell-free DNA and solid tumors revealed shared genomic alterations, providing orthogonal support for tumor origin and relatedness, while differences across analytes revealed genomic divergence and tumor heterogeneity.

**Conclusion:** AI-powered analysis leverages single-cell morphology for tumor cell identification and label-free enrichment, while integration with genomic profiling enables simultaneous interrogation of phenotypic and genomic heterogeneity in malignant effusions. This approach expands the potential of malignant effusions for liquid biopsy applications and biomarker development and discovery.

## INTRODUCTION

Malignant effusions are severe complications of advanced malignancy, characterized by fluid accumulation in the pleural cavity (pleural effusion) or the peritoneal cavity (ascites) ^1,2^. Pleural effusions most commonly occur in lung and breast cancers ^2–4^, whereas malignant ascites are frequently associated with ovarian, gastrointestinal, and breast cancers ^1,5^. Drainage of pleural or ascitic fluid by thoracentesis and paracentesis, respectively, is routinely performed for diagnostic evaluation and to relieve symptoms associated with fluid accumulation ^6,7^. Although the initial fluid collection is commonly evaluated by cytopathology to establish a diagnosis of malignancy, large volumes of fluid obtained during subsequent therapeutic procedures are often discarded ^8^. These repeatedly accessible specimens, which contain both intact metastatic tumor cells and cell-free DNA ^9^, represent an underutilized source of tumor material for liquid biopsy testing.

While tissue biopsy of metastatic lesions remains the clinical standard for biomarker testing ^10^, repeated sampling is often infeasible because of procedural risk and limited access to metastatic sites. These challenges have accelerated interest in liquid biopsy approaches that enable longitudinal characterization of the evolving genomic landscape of metastatic disease ^11,12^. Because drainage of fluids is clinically indicated, malignant effusions can serve as a source of metastatic cells for molecular profiling without requiring an additional invasive procedure ^13^. However, the tumor cell fraction (TCF) in malignant effusions can be highly variable, with tumor cells comprising only a small fraction of the cellular population amid abundant immune and other nonmalignant cells ^14,15^. The complex cellular admixture poses a major obstacle to isolating and molecularly characterizing tumor cells in these fluids. Existing approaches for tumor cell identification and enrichment from malignant effusions have relied on predefined molecular markers, including epithelial markers such as EpCAM and cytokeratins ^16–19^, or physical characteristics such as cell size ^18^. Although imaging-based approaches can also incorporate cellular morphology to identify tumor cells, these methods generally require fluorescent labeling and predefined molecular phenotypes ^16,17,19^. Such strategies may bias recovery toward tumor cells that conform to these predefined characteristics, potentially limiting representation of phenotypically and genotypically heterogeneous tumor populations in malignant effusions.

Advances in high-dimensional imaging and artificial intelligence (AI) offer an opportunity to use intrinsic cellular morphology as a label-free phenotype for cell identification and isolation ^20–22^. More recently, AI-powered morphology-based classification and enrichment have been applied to malignant effusions, demonstrating the feasibility of isolating tumor cells without relying on predefined molecular markers ^23^.

Whether such approaches can resolve morphological heterogeneity among tumor cells in malignant effusions and, importantly, whether morphologically identified tumor cells recapitulate the genomic features of the underlying malignancy remain incompletely understood. Establishing this connection between cellular phenotype and genomic identity would enable malignant effusions to be interrogated simultaneously as phenotypic and genomic representations of metastatic disease.

Here, we addressed these questions by applying AI-powered single-cell imaging and morphology analysis for label-free identification and enrichment of tumor cells (AI-identified TCs or AI-TCs) from malignant effusions. We evaluated AI-derived cell composition against conventional flow cytometry, identified phenotypically distinct tumor cell morphotypes through unsupervised analysis of image-derived features, and performed low-pass whole-genome and whole-exome sequencing of AI-TC-enriched fractions. By integrating morphological, copy-number, and mutational data with available matched solid tumor and effusion cell-free DNA profiles, including longitudinal samples, we investigated whether AI-TCs capture phenotypic heterogeneity and whether downstream genomic profiling provides orthogonal evidence of tumor origin, genomic relatedness, and divergence.

## METHODS

### Patients and samples

#### Patients

The study involved 10 patients with metastatic breast cancer and cytology-confirmed malignant pleural or ascitic fluid treated at the UCSF Comprehensive Cancer Center. Eligible participants enrolled in this non-therapeutic research study were identified by their primary breast oncologist and by the procedure team (UCSF Interventional Radiology Clinic) that performed the thoracentesis and paracentesis. Upon written informed consent, the patients were enrolled in the UCSF IRB-approved tissue collection protocol “Collection of Tumor Tissue, Blood, and Clinical Data from Patients with Breast Cancer” (CC10753).

#### Sample and data collection

We collected malignant effusion fluid for research after the initial visit, following diagnostic thoracentesis and/or paracentesis. Because these procedures are part of standard clinical care, and the drained fluid is typically discarded, participation in this study posed minimal additional risk to the participants. Serial malignant effusion fluid samples were collected when the participants returned for repeat malignant effusion drainage. Patient data (molecular and clinical) and research materials were de-identified and labeled with a unique study identification number.

#### Processing of malignant effusion samples

Pleural and ascitic fluid specimens were processed to generate single-cell suspensions for cryopreservation and molecular analyses (Fig. 1). The overall workflow consisted of (A) initial fluid processing and isolation of cellular material, (B) red blood cell (RBC) lysis when needed, (C) enzymatic digestion of cellular aggregates, (D) cryopreservation of isolated cells, and (E) processing and storage of cell-free supernatant.

**Figure 1.**
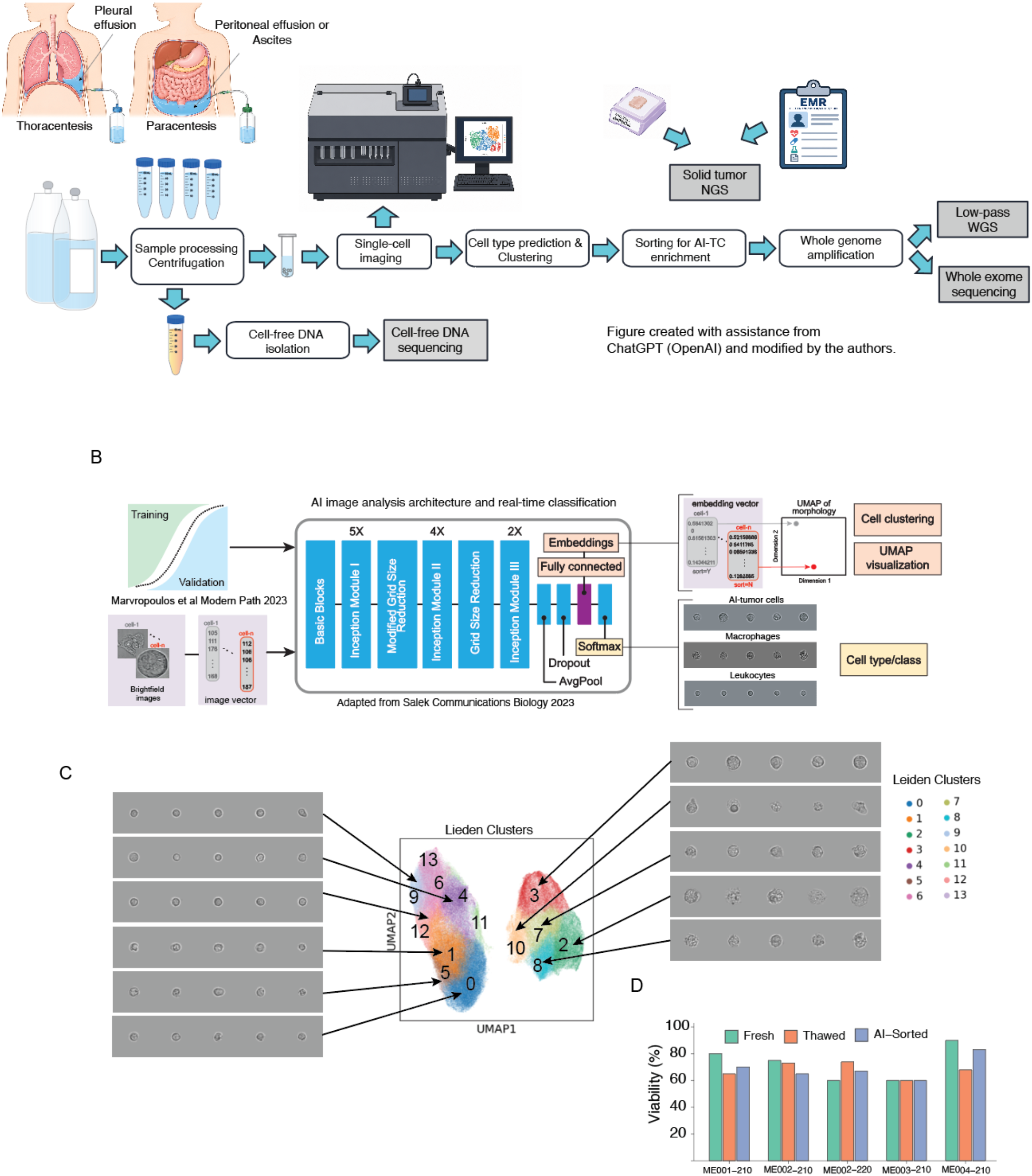
Experimental workflow for AI-powered morphological analysis of malignant effusions and downstream characterization. **A.** Overview of the workflow for processing malignant pleural effusion and ascites samples collected by thoracentesis or paracentesis, respectively. Following centrifugation and preparation of single-cell suspensions, aliquots were used for Deepcell imaging and sorting. Grey boxes indicate sequencing of tumor-derived analytes. Following sorting, AI-identified tumor cell (AI-TC)-enriched samples underwent whole-genome amplification and low-pass whole-genome sequencing and/or whole-exome sequencing. Cell-free DNA was isolated from the same fluids, and matched primary tumors were sequenced using the UCSF500 test. Available clinical next-generation tumor sequencing data were abstracted from patients’ medical records for comparison. **B.** Schematic of the AI image-analysis and deep-learning model architecture (malignant effusion AI model) used to generate image embeddings and classify cells. Brightfield cell images were transformed into image embeddings for cell-type prediction, dimensionality reduction, and clustering. **C.** Uniform manifold approximation and projection (UMAP) representation of unsupervised Leiden clusters with representative brightfield images illustrating cells from individual clusters. **D.** Cell viability was measured in fresh, thawed, and AI-sorted samples from five malignant effusion specimens.

##### Initial fluid processing

Pleural and ascitic fluid specimens, ranging from approximately 100 mL to several liters, were aliquoted into 50-mL conical tubes and centrifuged at 300 × g for 10 min at room temperature. The supernatant was retained and processed separately as described below.

Cell pellets were resuspended in 15 mL DMEM/F12 supplemented with 10% fetal bovine serum (FBS) and mechanically dissociated by repeated pipetting. Cell suspensions were passed through a 50-µm cell strainer. Material retained on the strainer was recovered by inverting the strainer onto a sterile Petri dish and flushing with 5 mL of phosphate-buffered saline (PBS). Recovered cellular aggregates were transferred to a separate 50-mL tube and subjected to enzymatic digestion as described below.

The filtrate was centrifuged at 300 × g for 10 min at room temperature. Samples with visible RBC contamination were subjected to RBC lysis before further processing (see next step). Cell pellets without substantial RBC contamination were resuspended in 3–5 mL DMEM/F12 supplemented with 10% FBS and combined into a single 50-mL tube. After enzymatic digestion of cellular aggregates, the resulting single-cell suspension was combined with the corresponding filtered cell fraction. The combined suspension was subsequently centrifuged at 300 × g for 10 min at room temperature.

##### Red blood cell lysis

For specimens with visible RBC contamination, RBC lysis was performed using RBC Lysis Buffer equilibrated to room temperature and protected from light. A 1× working solution was prepared by diluting 10× RBC Lysis Buffer 1:10 in deionized water. Cell pellets were resuspended in 10 mL of 1× RBC Lysis Buffer and incubated for 15 min at room temperature, protected from light. Samples were then centrifuged at 300 × g for 5 min.

RBC lysis was repeated as necessary, for up to three cycles, until visible RBC contamination was eliminated. Following the final wash, cell pellets were resuspended in 10 mL DMEM/F12. Total cell number and cell viability were determined before cryopreservation.

##### Enzymatic digestion of cellular aggregates

Cellular aggregates retained during filtration were enzymatically dissociated using collagenase. Lyophilized collagenase (50 mg) was reconstituted in 50 mL PBS containing 0.5% bovine serum albumin (BSA) to generate a 1 mg/mL stock solution. Reconstituted collagenase was aliquoted and stored at −20°C until use.

For digestion, 1 mL of the 1 mg/mL collagenase solution was added to the recovered cellular aggregates, and the total volume was adjusted to 10 mL with PBS. Samples were incubated at 37°C with agitation at 300 rpm and examined microscopically at approximately 10-min intervals. Digestion was terminated when a predominantly single-cell suspension was obtained, with digestion times of up to 90 min depending on the size and extent of the cellular aggregates.

Following digestion, samples were centrifuged at 300 × g for 10 min at room temperature. The supernatant was removed, and the cell pellet was resuspended in approximately 5 mL DMEM/F12 supplemented with FBS, with the volume adjusted as necessary according to pellet size. The resulting suspension was passed through a 50-µm cell strainer and combined with the corresponding cell fraction obtained during initial fluid processing.

##### Cell cryopreservation

Single-cell suspensions were centrifuged at 300 × g for 5 min at room temperature, and the supernatant was carefully removed. Cell pellets were resuspended in chilled (2–8°C) CryoStor CS10 cryopreservation medium at a target concentration of 1–2 × 10^6 cells/mL. Aliquots of 1 mL were transferred to cryovials and incubated at 2–8°C for 10 min.

Cells were cryopreserved using controlled-rate cooling at approximately −1°C/min or an isopropanol-based freezing container. Cryovials were subsequently transferred to liquid nitrogen storage (≤−135°C) the following day and maintained under cryogenic conditions until further analysis.

##### Processing of cell-free effusion supernatant

Following the initial centrifugation of pleural or ascitic fluid, an aliquot of the recovered supernatant was reserved for cell-free DNA extraction and sequencing. The remaining supernatant was transferred to 50-mL conical tubes and centrifuged at 16,000 × g for 10 min at 4°C to remove residual cellular material and debris. The clarified supernatant was aliquoted into 4-mL cryovials and stored at −80°C until subsequent analyses.

### Cell assays

#### Flow cytometry

The distributions of different cell populations in malignant effusions were estimated using standard flow cytometry procedures. Fluorescently labeled antibodies to CD45, CD68, EpCAM, and Claudin-4 were added to fixed single*-*cell suspension samples. Flow cytometry was performed on a BD FACSMelody machine using the following gating strategy to identify cell types: (1) leukocytes were identified by CD45 expression; (2) macrophages, a leukocyte subset, were identified by the co-expression of CD45 and CD68; and (3) putative tumor cells were identified by EpCAM and/or Claudin-4 ^24–26^ expression and the lack of CD45 expression. The gating strategy enabled estimation of the TCF and the proportions of other cell types in each sample.

#### Cell imaging, classification (embedding) analysis, and cell sorting using the Deepcell platform

The Deepcell instrument consists of hardware (microfluidic chip, high-resolution imager, and real-time signal processor), a deep learning convolutional neural network (malignant effusion AI model), and proprietary software for image analysis.

##### Cell imaging

Single cells in suspension were hydrodynamically and inertially focused within a microfluidic cartridge and passed through the imaging region of the instrument. High-speed, high-resolution brightfield imaging was performed on individual cells in flow. The captured brightfield images were cropped around the cell (299 × 299 pixels), and the resized images were then fed in real time into a convolutional neural network, called the “malignant effusion AI model,” to extract computer-vision-derived multidimensional morphological and deep-learning features for cell-type classification.

##### Imaging analysis and cell classification

The convolutional neural network (malignant effusion AI model) was developed using a modified InceptionV3 architecture with 48 layers and approximately 24 million parameters. The model was trained on images of highly pure populations of breast tumor cells and immune cells, including leukocytes and macrophages. The training and test datasets consisted of ∼2.6 million images and ∼2.0 million images, respectively ^23^.

The convolutional neural network converted each cell image into an embedding vector that provides a quantitative representation of cellular morphology. The representation incorporated 51 human-interpretable morphometric features (e.g., cell size, shape, and texture) and 64 deep-learning-derived features that capture additional morphological patterns not readily interpretable by human observers ^27^. After generating the embedding vector, the final softmax layer of the convolutional neural network produced class probabilities for each cell type (classification): tumor cells, leukocytes (non-macrophage), macrophages, and “other” cells. Each cell was assigned to the class corresponding to the highest predicted probability using the argmax function.

The TCF in each sample was calculated by dividing the number of cells predicted as tumor cells by the total number of cells with predictions. In a previous study, the model’s classifier demonstrated excellent discrimination between breast carcinoma and non-carcinoma cells, with an area under the receiver operating characteristic curve (AUC) of 0.96 ^23^.

##### Cell sorting

Each cell was assigned a class in real time as it traveled through the microfluidic cartridge. The classification then triggered a decision that routed the cell to the appropriate collection reservoir. This was performed by the signal processor, which converted the classification into precisely timed hardware commands, ensuring the correct cell was sorted. If the classification probability, as determined by the softmax activation function, exceeded 0.85, pneumatic sorting valves were activated to direct the cell to a specific collection reservoir. Cells below the probability threshold or belonging to other classes were routed to the waste reservoir. The sorted target cells were then retrieved by pipetting them from the collection reservoir and transferred to an Eppendorf tube for cell lysis and whole-genome amplification.

##### Data visualization

The proprietary image-analysis software (Axon) enabled visualization of single-cell images and analysis of cell populations in each sample ^28^. Cells were projected into two-dimensional morphology space using Uniform Manifold Approximation and Projection (UMAP) ^29^. Unsupervised Leiden clustering ^30^ was performed on the high-dimensional morphological representations to identify groups of cells with similar morphological phenotypes (morphotypes). Leiden cluster assignments were overlaid on the UMAP for visualization. Cell-density plots were generated in the common UMAP space to visualize regions preferentially occupied by cells from individual samples or cell populations.

##### Feature importance analysis

Random forest classifiers were used to identify morphological and deep-learning-derived features associated with each morphotype cluster. A one-cluster-versus-all approach was used, in which cells assigned to each morphotype cluster were classified against cells from all other clusters. Features were ranked according to their importance scores, with higher scores indicating greater contribution to distinguishing a given morphotype cluster from the remaining clusters.

Imaging analysis and subsequent next-generation sequencing of AI-TC-enriched fractions (see next section) were performed as described below (Table S1, Table S2).

#### Cell viability

For viability assessment, cells were stained with trypan blue, and viable cells were counted under a microscope. Cell viability was assessed in freshly prepared single-cell suspensions, cells thawed after cryopreservation, and cells sorted using the Deepcell instrument.

### Sequencing of AI-identified tumor cells and normal control cells

#### Whole-genome amplification, library preparation, and sequencing

Whole-genome amplification was performed using ResolveDNA WGA (BioSkryb Genomics) on genomic DNA released after cell lysis, according to the manufacturer’s protocol.

Libraries for copy-number analysis were constructed from 500 ng of WGA product using the Watchmaker DNA Library Prep Kit with Fragmentation (Watchmaker Genomics). For low-pass whole-genome sequencing, the libraries were sequenced to <1X coverage on an Illumina NextSeq 1000 (Illumina) instrument with NextSeq1000/2000 P2 Reagents (300 Cycles) v3 (Illumina).

#### Low-pass whole-genome sequencing for copy-number analysis

Copy-number alterations were analyzed from low-pass whole-genome sequencing data using ichorCNA software (bioconda r-ichorcna version 0.3.2) ^31^. Briefly, the raw FASTQ files were aligned to the human reference genome, GRCh38.p13, using the BWA-MEM algorithm to generate BAM files ^32^. Read-count coverage was calculated from BAM files using the readCounter command in HMMcopy with a bin size of 500 kb. The read counts were used as inputs to the ichorCNA software for copy-number analysis. Genome-wide copy-number profiles and segmented copy-number calls were generated using the optimal ichorCNA solution. Tumor fraction (TF) was estimated using ichorCNA lower-bound estimates set to 3% for visualization and calculation. Tumor-fraction estimates <3% were considered below the previously established lower limit for reliable tumor detection by ichorCNA ^31^.

#### Whole-exome sequencing (WES) of AI-TC-enriched and normal control cells

Sequencing libraries for AI-TC-enriched (tumor) and normal control cells were prepared from 500 ng of WGA product using Watchmaker library preparation reagents. Normal control libraries were generated from leukocytes, except for patients ME003, ME007, and ME010, for whom macrophages served as normal controls. Leukocytes (CD45+) and macrophages (CD45+/CD68+) were isolated from the same sample by fluorescence-activated cell sorting (FACS) performed in duplicate.

Exome capture was performed using 187.5 ng of library per sample with the Twist Human Core Exome Panel v2.1. The libraries were sequenced to a median target coverage of ∼260x. Paired-end Illumina sequencing reads were aligned to the GRCh38 human reference genome using BWA-MEM. Alignments with mapping quality scores <20 were removed using SAMtools, and PCR and optical duplicates were identified using Picard MarkDuplicates. Base quality score recalibration was performed using GATK ApplyBQSR with gnomAD hg38 known sites and restricted to the Twist Exome target regions.

Somatic variants were called with GATK Mutect2 (v4.2.2.0), restricted to Twist Exome 2.1 target intervals on the GRCh38 no-alt reference. Variant calling was performed in multi-sample tumor–normal mode, jointly analyzing sequences from two replicate libraries of each AI-TC-enriched sample against two matched normal control libraries (see previous section). The same normal control sequences served as the reference for all timepoints from a given patient. The GATK 1000 Genomes panel of normals and the gnomAD allele-frequency-only germline resource were used to exclude recurrent technical artifacts and germline variants. Calls were filtered with FilterMutectCalls (v4.2.2.0) using the default optimal F-score threshold; contamination and read-orientation bias models were not applied. Single-nucleotide variants were extracted with bcftools (v1.14).

To complement the GATK Mutect2 variant calls, two additional somatic variant callers were applied to the recalibrated BAM files from the AI-TC-enriched and normal control cells. Strelka2 ^33^ was run through the nf-core/Sarek pipeline (v3.6.0) ^34^ in whole-exome sequencing mode using the GATK GRCh38 reference genome. Strelka SNV and indel VCFs were concatenated per sample using bcftools ^35^.

VarScan (v2.4.6) ^36^ was run independently. Pileup files were generated using samtools mpileup (v1.21) ^35^ with a minimum mapping quality of 1, restricted to exome target regions. VarScan somatic calling was performed with a minimum coverage of 3 and a minimum variant frequency of 0.01.

Caller-specific quality filters were applied prior to consensus generation: 1) MuTect2: Retained variants passing GATK FilterMutectCalls (PASS filter). 2) Strelka2: Retained PASS-only variants. VAF was calculated from tier 1 allele counts. VarScan: Retained PASS-only somatic variants (somatic status SS = 2) with somatic confidence score ≥ 20, tumor VAF ≥ 0.05, total depth ≥ 10, alt-supporting reads ≥ 3, and normal alt-supporting reads ≤ 1 to remove germline leakage.

For all callers, somatic tumor variants with VAF <0.05 were excluded based on the established limit of detection for whole-exome sequencing ^37^. For patients with multiple samples from a single timepoint, VCFs were merged using bcftools, and the mean allele frequency across tumor samples was computed for each variant.

A combined variant set was generated for each patient-timepoint by taking the union of somatic variants passing quality filters across all three callers. Variants were annotated using vcf2maf (v1.6.22) (mskcc/vcf2maf: vcf2maf v1.6.16, n.d.) with Ensembl VEP ^38^, selecting a single effect per variant based on the Ensembl canonical transcript. The reference genome used was GRCh38.p13 with the GRCh38 VEP cache.

Combined heatmaps of somatic variants were generated using ComplexHeatmap ^39^. Variant heatmaps displayed the top 20 most frequently mutated genes, restricted to coding variant classifications (missense, nonsense, frameshift, in-frame indels, splice site, and translation start site mutations). Clinical annotations included effusion source, timepoint, and patient identity. Cancer-associated gene oncoplots used oncogenic gene lists from the maftools package ^40^, supplemented with *ESR1* and *ESR2*. TF was estimated from the WES data using ichorCNA, with analysis restricted to genomic regions targeted by the Twist Human Core Exome Panel v2.1.

### Sequencing of cell-free DNA and solid tumors

#### Cell-free DNA isolation

Fluids from effusion samples collected in Cell-Free DNA BCT tubes were subjected to sequential centrifugation at 1,900 × g for 10 min at room temperature and 16,000 × g for 10 min at 4°C to remove cellular debris, yielding approximately 5.2 mL of clarified supernatant. Samples were treated with proteinase K, incubated at 60°C for 20 min, cooled on ice for 5 min, and cell-free DNA was isolated using the MagMAX Cell-Free DNA Isolation Kit on a KingFisher Apex instrument. Purified DNA was assessed using an Agilent Fragment Analyzer, with cell-free DNA and high-molecular-weight DNA defined as fragments of 100-1,000 bp and 1,000-6,000 bp, respectively, and total DNA yield was quantified using the Qubit dsDNA High Sensitivity Assay.

#### Primary tumors

Formalin-fixed, paraffin-embedded tumor blocks were cut into 5-10 unstained 10-µm sections. Regions with tumor cellularity greater than 25% were identified by histologic review using an adjacent hematoxylin and eosin (H&E)-stained slide as a guide. Genomic DNA was isolated from macrodissected tissue samples using the QIAamp DNA FFPE Tissue Kit (Qiagen).

#### Targeted sequencing and copy-number analysis using the UCSF500

Sequencing of cell-free DNA and primary tumor DNA was performed using the research use only (RUO) pipeline of the UCSF500 Cancer Gene Panel test provided on a fee-for-service basis by the UCSF Clinical Cancer Genomics Laboratory. The UCSF500 is a hybrid-capture next-generation sequencing assay targeting the coding regions of cancer-associated genes, selected intronic regions of genes recurrently involved in structural rearrangements, and genomic regions distributed across the chromosomes to assess copy-number alterations. The current assay targets 529 cancer-associated genes and selected introns of 47 genes. The sequencing libraries were enriched by hybrid-capture using custom oligonucleotide probes and sequenced as paired-end reads on an Illumina NovaSeq 6000. The sequence reads were computationally deduplicated and aligned to the human reference genome (UCSC hg19/NCBI build 37).

For variant analysis, single-nucleotide variants (SNVs) and small insertions and deletions (indels) were identified using the UCSF500 bioinformatics pipeline. The UCSF500 pipeline uses FreeBayes, GATK variant callers, and Pindel for SNV and indel detection, with Delly and Pindel for detecting larger indels and structural variants. Variants were annotated using ANNOVAR, and candidate SNVs, indels, and structural variants were reviewed using the Integrative Genomics Viewer (IGV). Germline polymorphisms and recurrent sequencing artifacts were removed through database- and pipeline-based filtering.

Genome-wide copy-number and zygosity analysis was performed from UCSF500 sequencing data using CNVkit. The UCSF500 panel includes genomic targets distributed at intervals across the genome, enabling assessment of chromosome-level and focal copy-number alterations, as well as alterations affecting the targeted cancer genes. CNVkit uses sequencing coverage from targeted regions, together with additional sequencing information available across the capture assay, to estimate relative copy number.

#### Clinical sequencing of solid tumors

Results from clinically performed next-generation sequencing of solid tumor specimens were obtained by retrospective review of the electronic medical records. Molecular profiling had been performed as part of routine clinical care and was not conducted specifically for this study. Available clinical molecular pathology reports were reviewed, and reported somatic genomic alterations were abstracted for comparison with genomic profiles generated from malignant effusion tumor cells and effusion cell-free DNA. When available, the anatomic site and date of tumor sampling, sequencing assay, and date of molecular testing were recorded.

## RESULTS

### Patients, samples, and assays

The study included 10 patients with metastatic breast cancer who developed malignant effusions, including seven with hormone receptor-positive/HER2-negative disease and three with triple-negative breast cancer; none had HER2-positive disease (Table S1). Eight patients had pleural effusions, and two had ascites. The cohort was predominantly female, with six patients identifying as White. Ages ranged from 47 to 80 years, with a mean age of 61 years. Serial effusion samples were available from six patients, with 2–7 samples collected per patient.

Pleural and ascitic fluids obtained by therapeutic thoracentesis or paracentesis were processed to generate single-cell suspensions and cell-free supernatants (Fig. 1A). Single-cell suspensions were cryopreserved and subsequently analyzed by AI-powered single-cell imaging (Deepcell) and flow cytometry. Matched cell-free DNA (n = 17 samples) from the same fluids and archival tumor blocks (n = 4 patients) were collected for sequencing (Fig. 1A). Data from clinical sequencing of tumors (n = 6 patients) were extracted from patients’ medical records. The assays performed for individual samples are summarized in Table S2.

### AI-powered single-cell morphology enables label-free cell-type classification

We applied the Deepcell platform to perform high-throughput, label-free morphological analysis of single cells from malignant effusions. As cells flowed through the microfluidic system, brightfield images were acquired and analyzed in real time using a previously developed convolutional neural network trained to distinguish tumor cells from other cell populations in malignant effusions ^23^ (Fig. 1B; Fig. S1A). The model generated morphological embeddings comprising 51 interpretable morphometric features, including cell size, shape, intensity, and texture, and 64 deep-learning-derived features learned directly from cell images (Fig. S1B). These quantitative representations enabled visualization of cells in a 2-dimensional space. Fig. 1C shows the results of unsupervised clustering of all imaged cells (n = 25 samples) using the Leiden algorithm, along with representative brightfield images from each cluster.

### Cell viability is preserved following imaging and sorting

Because genomic profiling requires recovery of intact cells, we evaluated cell viability before and after processing with the Deepcell instrument. Viability was assessed in fresh samples, following cryopreservation and thawing, and after imaging and cell sorting. Across the five samples evaluated, cell viability remained comparable after processing (Fig. 1D). Together with previous reports demonstrating preservation of cell viability following imaging and sorting ^20,23^, these findings support recovery of intact cells suitable for downstream molecular analyses.

### AI-powered single-cell imaging reveals heterogeneous cellular composition of malignant effusions

To characterize the cellular composition of malignant effusions, we applied AI-powered cell classification across 25 samples from 10 patients. Visualization of morphological embeddings using UMAP revealed two major clusters, with leukocytes (non-macrophage) predominantly occupying one cluster and macrophages and AI-TCs occupying a second (Fig. 2A). Although AI-TCs and macrophages showed partial overlap in several individual morphometric features, including cell size-related measures, as previously observed ^23^ (Fig. S1C), their separation within the multidimensional morphological representation indicates that classification incorporates morphological information beyond cell size alone.

**Figure 2.**
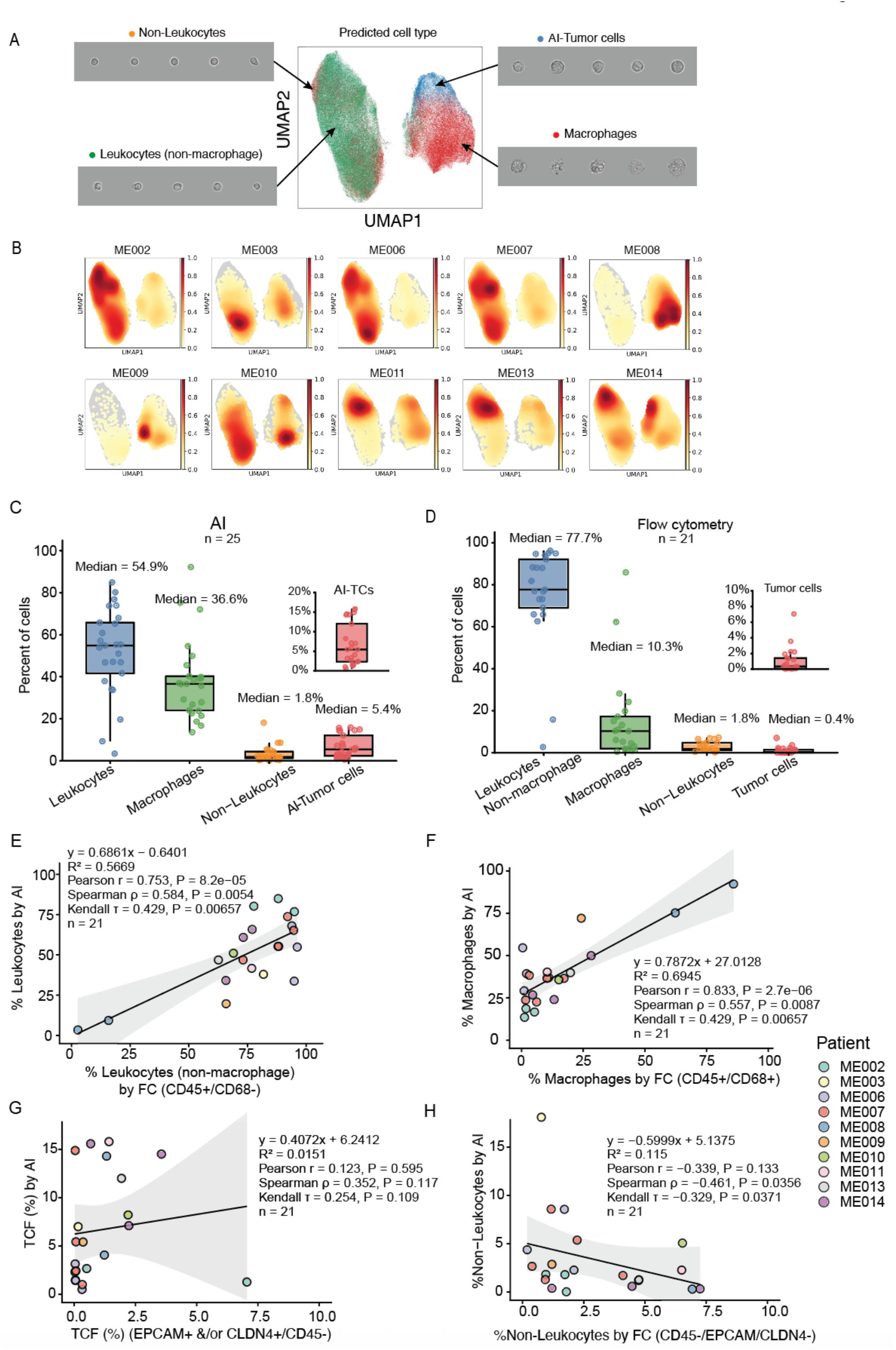
AI-powered single-cell imaging characterizes cellular composition of malignant effusions and correlates with flow cytometry for major immune-cell populations. **A.** Uniform manifold approximation and projection (UMAP) of single cells from malignant effusion samples based on features extracted from brightfield images and colored by predicted cell type. Representative single-cell images are shown for non-leukocytes, leukocytes (non-macrophage), AI-identified tumor cells (AI-TCs), and macrophages. **B.** UMAP density plots showing the distribution of cells from individual patients within the AI-derived feature space. **C.** Cellular composition of malignant effusion samples determined by AI-powered single-cell classification. Box plots show the proportions of cells classified as leukocytes (non-macrophage), macrophages, non-leukocytes, and AI-TCs across samples (enlarged in the inset). Median percentages are indicated. **D.** Cellular composition of matched malignant effusion samples determined by flow cytometry. Box plots show the proportions of leukocytes (non-macrophage), macrophages, non-leukocytes, and tumor cells (enlarged in the inset). Median percentages are indicated. **E–H.** Comparison of cell-type proportions determined by AI-powered classification and flow cytometry in matched samples: **E.** leukocytes (non-macrophage), **F.** macrophages, **G.** tumor cells (TCF-tumor cell fraction), and **H.** non-leukocytes. Points represent individual samples and are colored by patient. Solid lines indicate linear regression fits with 95% confidence intervals. Regression equations, R², Pearson, Spearman, and Kendall correlation coefficients with corresponding *P* values, and sample sizes are shown.

Cellular composition varied among patients, as demonstrated by sample density distributions projected onto a common UMAP representation (Fig. 2B). Leukocytes were the predominant cell population overall (median, 54.9%; range, 3.4–85%), followed by macrophages (median, 36.6%; range, 13.5–92.3%), except in serial samples from patient ME008, where macrophages were the most abundant (Fig. S2B). Non-leukocytes represented a smaller fraction of the cell population (median, 1.8%; range, 0.03–18.1%; Fig. 2C). Consistent with the malignant diagnosis by cytopathology, AI-TCs were detected in all samples (median, 5.4%; range, 0.51–15.8%; see Fig. 2C inset). Serial samples demonstrated changes in cellular composition within individual patients over time (Fig. S2A), indicating both intra- and inter-patient heterogeneity.

### AI-derived estimates of major immune-cell populations correlate with flow cytometry

AI-derived estimates of the major immune-cell populations correlated significantly with flow cytometry measurements (n = 21 paired samples, Fig. 2D). Leukocyte proportions measured by AI correlated with CD45+/CD68− leukocytes by flow cytometry (Pearson r = 0.75, P = 8.2 × 10⁻⁵; Fig. 2E). AI-derived macrophage proportions also correlated with CD45+/CD68+ macrophages (Pearson r = 0.69, P = 2.7 × 10⁻⁶; Fig. 2F), including the marked macrophage-predominant effusions in patient ME008 (Fig. S2C), which was also observed using AI-derived cell classification (Fig. S2B, previous section). In contrast, estimates of TCF did not significantly correlate between the two approaches (Pearson r = 0.12, P = 0.595, Fig. 2G), whereas non-leukocyte populations were negatively correlated (Pearson r = −0.34, P = 0.133; Fig. 2H). Although initial AI-based cell classification used the highest predicted probability, a minimum confidence threshold of 0.85 was subsequently adopted for cell sorting. In a post hoc exploratory analysis, increasing the confidence threshold reduced AI-derived tumor-cell fraction (TCF) estimates but improved their correlation with flow cytometry (Fig. S2D). Among 21 samples, Spearman correlation increased from ρ = 0.33 (P = 0.14) when all AI tumor-cell calls were included, regardless of classifier confidence, to ρ = 0.47 (P = 0.034) when only calls with confidence ≥0.95 were included.

### Unsupervised analysis identifies distinct tumor cell morphotypes in malignant effusions

To determine whether AI-TCs exhibited morphological heterogeneity, we performed unsupervised clustering of their morphological embeddings using the Leiden algorithm. This analysis identified four distinct clusters, hereafter referred to as “morphotypes**”** (Fig. 3A). The distribution of morphotypes varied substantially among patients (Fig. 3B, Fig. 3C), and serial samples from individual patients also showed changes in morphotype composition over time (Fig. S3A, Fig. S3B), demonstrating both intra- and inter-patient morphological heterogeneity.

**Figure 3.**
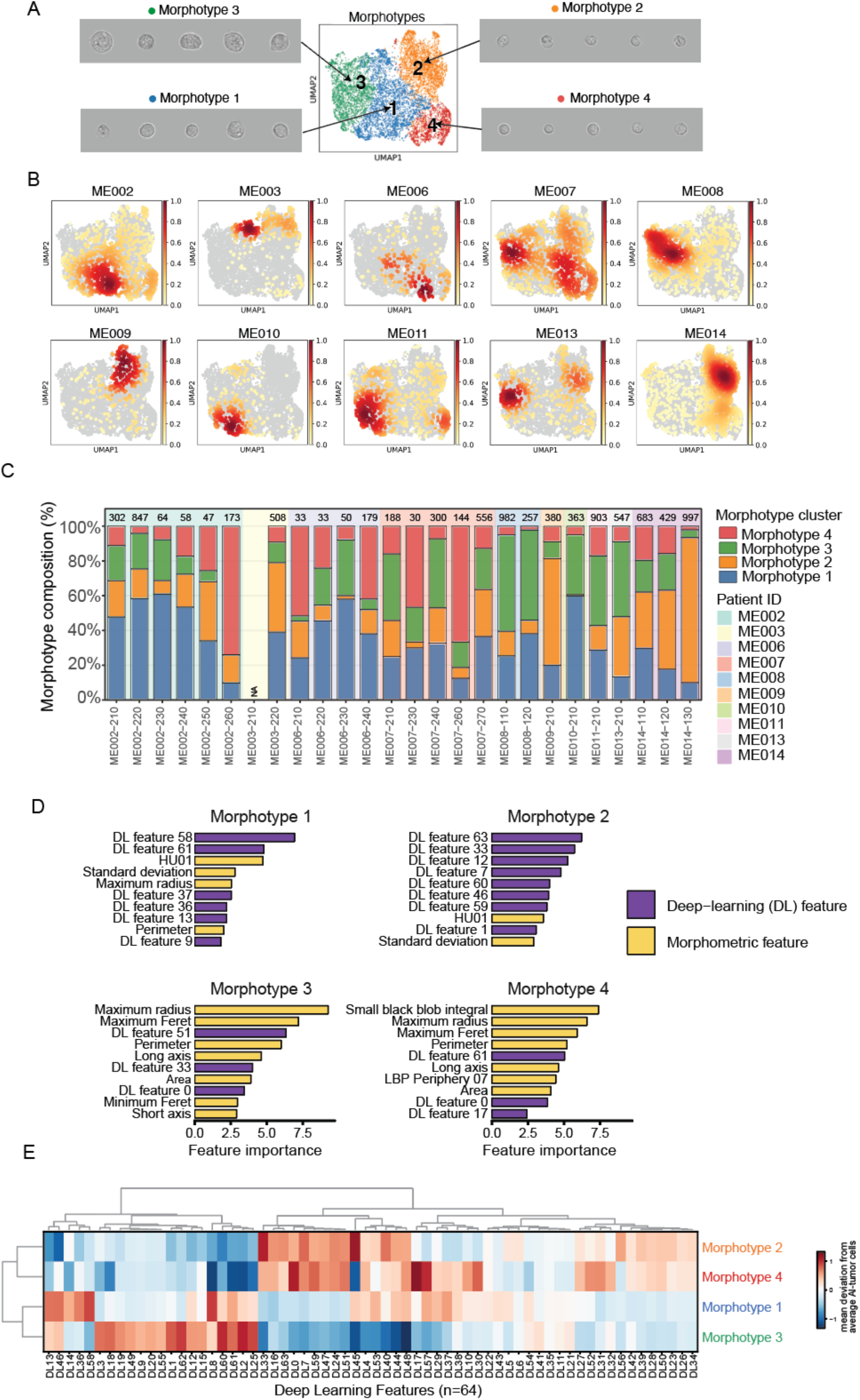
Unsupervised clustering identifies distinct AI-identified tumor cell morphotypes in malignant effusions. **A.** Uniform manifold approximation and projection (UMAP) representation of AI-identified tumor cells (AI-TCs) showing four clusters identified by unsupervised clustering, designated Morphotypes 1–4. Representative brightfield images illustrate cells from each morphotype. **B.** Patient-specific density distributions of AI-TCs in UMAP space for 10 patients, illustrating inter-patient variation in morphotype distributions. **C.** Morphotype composition of individual samples. Stacked bars show the proportions of morphotype clusters 1–4; numbers above bars indicate the number of AI-identified tumor cells analyzed, and background shading identifies patients. **D.** One-versus-all random forest feature-importance analysis identifying the features that distinguish each morphotype from the remaining morphotypes. The 10 most important features for each morphotype are shown and classified as deep-learning (DL) or morphometric features. **E.** Hierarchically clustered heatmap of 64 deep-learning image features across the four morphotypes. Values represent the mean deviation of each feature from the average AI-TC feature value.

To identify the morphological and deep-learning features that distinguish each morphotype, we performed one-versus-all random forest analyses (Table S3). Morphotypes 1 and 2 were distinguished predominantly by deep-learning-derived features, whereas Morphotypes 3 and 4 showed greater contributions from interpretable morphometric features (Fig. 3D). Morphotype 3 was characterized by larger cell-size and shape-related measurements, including perimeter, area, maximum radius, maximum Feret diameter, and long-axis length, whereas morphotype 4 was associated with smaller values for these features and showed a strong contribution from the texture-related small-black-blob integral feature (Fig. S3C). Table S4 lists definitions for the selected morphological features.

Analysis of deep-learning feature patterns further showed distinct, non-human-interpretable signatures among the four morphotypes (Fig. 3E). These morphotype-associated signatures were also observed across samples from different patients, with tumor cells assigned to the same morphotype showing similar deep-learning feature profiles regardless of patient origin (Fig. S3D). Together, our analysis demonstrates that AI-TCs in malignant effusions can constitute morphological subpopulations that vary both across patients and over time within individual patients.

### Morphology-guided enrichment enables genomic copy-number profiling of tumor cells from malignant effusions

To determine whether morphology-guided sorting could enrich tumor cells for genomic characterization, we subjected AI-TC-enriched and matched unenriched cell samples (immune-cell-predominant) to low-pass whole-genome sequencing following whole-genome amplification (see Methods). TF was estimated from genome-wide copy-number profiles using ichorCNA. Among paired samples, AI-TC enrichment increased sequencing-derived TFs in 14 of 26 samples (mean, 3-fold; range, 1–15.5-fold), including at least one sample from 7 of 10 patients (Fig. 4A; Fig. S4A).

**Figure 4.**
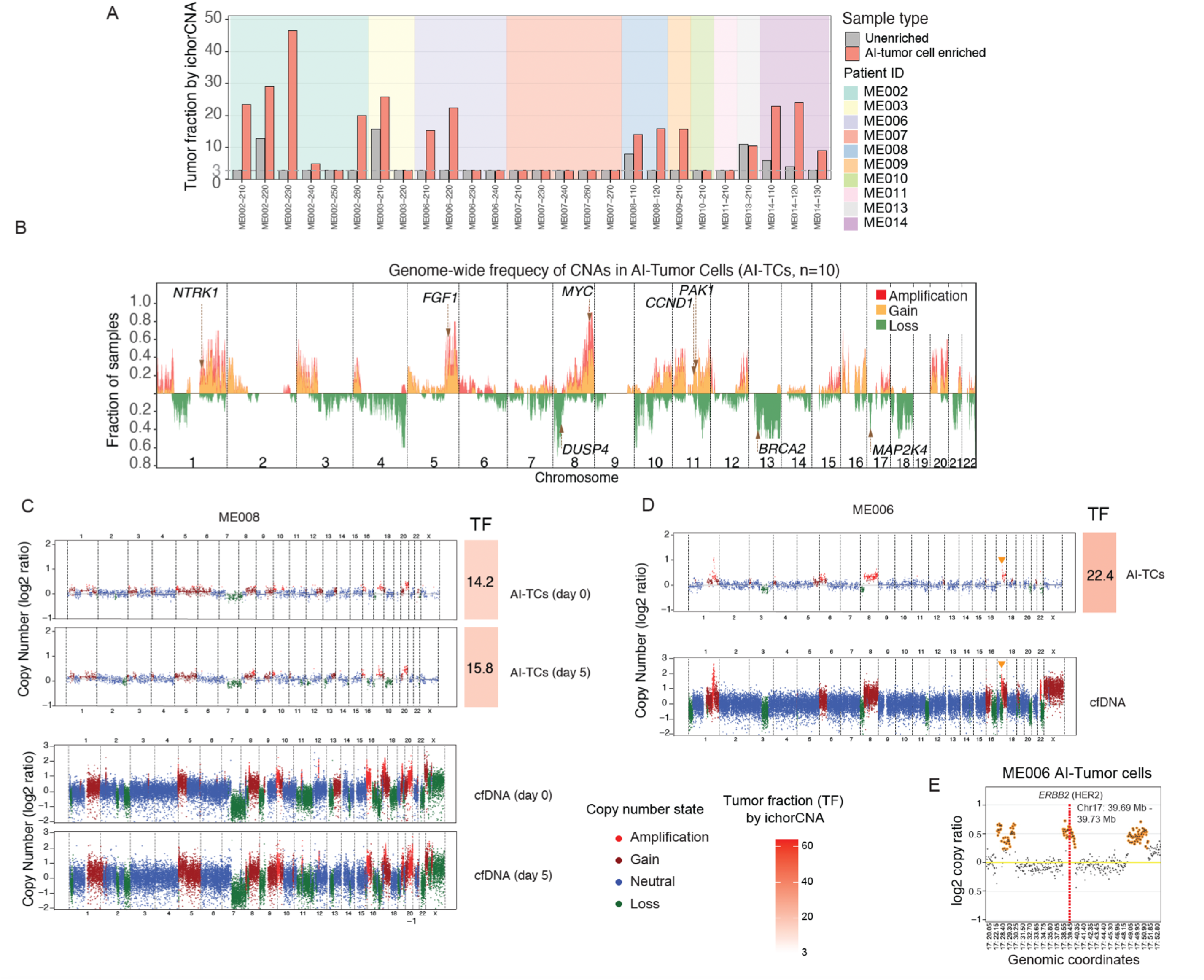
Morphology-guided enrichment enables copy-number profiling of AI-identified tumor cells from malignant effusions. **A.** Tumor fraction (TF) estimated by ichorCNA from low-pass whole-genome sequencing of paired AI-identified tumor cell (AI-TC)-enriched and unenriched (immune-cell-dominant) samples. Bars represent individual samples and are grouped by patient. The lower limit of TF estimation by ichorCNA was 3% ^31^. **B.** Genome-wide frequency of copy-number alterations detected by low-pass whole-genome sequencing in AI-TC-enriched samples from 10 patients. The fraction of samples harboring amplification, gain, or loss is plotted across chromosomes 1–22. Selected cancer-associated genes located within recurrently altered regions are indicated. **C.** Genome-wide copy-number profiles of serial AI-TC-enriched fractions and matched effusion cell-free DNA from patient ME008. AI-TC samples and matched cell-free DNA were collected at days 0 and 5. **D.** Genome-wide copy-number profiles of an AI-TC-enriched fraction and matched effusion cell-free DNA from patient ME006. Arrow indicates focal amplification on 17q. **E.** Detailed copy-number profile of chromosome 17 in an AI-TC-enriched sample from patient ME006, whose breast cancer was clinically classified as HER2-negative, demonstrating focal amplification encompassing *ERBB2*, which encodes HER2. The genomic location of *ERBB2* is indicated. The corresponding amplification was also detected in matched effusion cell-free DNA in (D). In (C–D), copy-number states are indicated as amplification, gain, neutral, and loss. TF estimated by ichorCNA is shown for the AI-TC-enriched samples.

Copy-number profiles varied substantially among patients, demonstrating inter-patient genomic heterogeneity. Comparison with matched unenriched fractions generally showed more pronounced copy-number alteration signals following AI-TC enrichment (Fig. S4B). Serial unenriched samples from patient ME002 similarly showed weak or absent copy-number alteration signals at several timepoints, with low ichorCNA-estimated TF, consistent with immune-cell-dominant samples with non-aberrant genomes (Fig. S4C). Interestingly, we did not observe copy-number alterations in ME010 or in longitudinal samples from ME007.

Genome-wide copy-number profiling of AI-TC-enriched samples from 10 patients identified recurrent chromosomal gains and losses characteristic of breast cancer, including gains of 1q and 8q and loss of 8p (Fig. 4B). Focal alterations included amplifications encompassing cancer-associated genes such as *MYC* at 8q24.21 and *CCND1* at 11q13.3, as well as losses involving regions containing *DUSP4* at 8p12 and *BRCA2* at 13q13.1 (Fig. 4B). Known oncogenes or tumor suppressor genes located on chromosome regions with gains or losses, respectively, are listed in Table S5.

To determine whether copy-number alterations detected in AI-TCs were also represented in an independent tumor-derived analyte, we compared AI-TC profiles with matched effusion cell-free DNA. In patient ME008, AI-TC-enriched fractions from two ascites samples collected five days apart showed similar genome-wide copy-number alteration patterns, many of which were also evident in matched serial cell-free DNA samples (Fig. 4C). Similarly, the AI-TC-enriched fraction from patient ME006 showed multiple copy-number alterations that were also observed in matched effusion cell-free DNA (Fig. 4D). Notably, although ME006 had clinically HER2-negative disease, focal amplification of the *ERBB2* locus, which encodes HER2, was detected in the AI-TC-enriched fraction (Fig. 4E). Amplification of the corresponding *ERBB2* region was also evident in matched effusion cell-free DNA (log2 copy ratio, 1.2; Fig. 4D). Clinical testing by fluorescence in situ hybridization (FISH) of tumor cells obtained from fine needle aspirate of pleural fluid also revealed *ERBB2/HER2* amplification (average ratio of HER2: Centromere 17, 2.6). Together, these tests provide independent support for this alteration. These findings demonstrate that morphology-guided enrichment enables profiling of genome-wide copy-number alterations in AI-TC-enriched samples from malignant effusions and recovers tumor-associated alterations also represented in matched effusion cell-free DNA.

### Whole-exome sequencing of AI-TC-enriched samples identifies cancer-associated mutations and genomic relatedness across tumor-derived analytes

The same whole-genome-amplified DNA from AI-TC-enriched samples used for low-pass whole-genome sequencing was subsequently analyzed by whole-exome sequencing (WES). Across the cohort, WES identified somatic variants in multiple cancer-associated genes, including *PIK3CA, ESR1, KMT2D,* and *CREBBP*, among others (Fig. 5A). Mutation profiles varied across patients and serial samples, demonstrating substantial inter-patient and intra-patient genomic heterogeneity (Fig. 5A; Fig. S5, Table S6). Detailed longitudinal mutation profiles from patients with serially collected malignant effusions are shown in Fig. S5.

**Figure 5.**
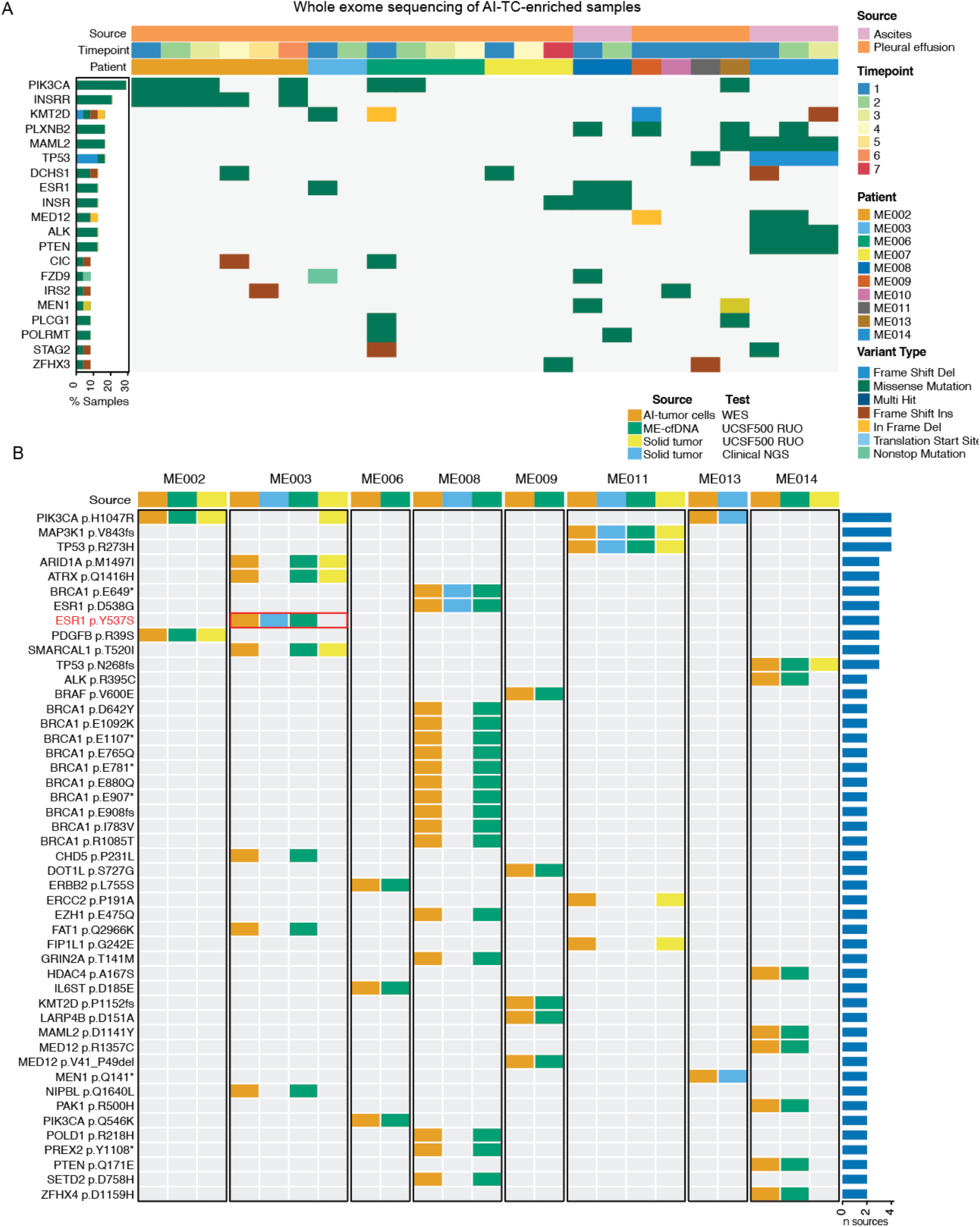
Whole-exome sequencing of AI-identified tumor cells reveals cancer-associated mutations and genomic relatedness across tumor-derived analytes. **A.** Oncoplot summarizing recurrent somatic variants detected by whole-exome sequencing of AI-TC-enriched malignant effusion samples. Columns represent individual samples and are annotated by fluid source, collection timepoint, and patient. Rows represent recurrently altered genes, with tile color indicating variant type. The percentage of samples harboring alterations in each gene is shown at left. **B.** Comparison of somatic variants detected by whole-exome sequencing of AI-TC-enriched samples with variants identified in available matched malignant-effusion cell-free DNA and solid-tumor samples. Columns are grouped by patient and represent available genomic data sources, including AI-TCs analyzed by whole-exome sequencing, effusion cell-free DNA, and solid tumors analyzed using the Researcher Use Only (RUO) pipeline of the UCSF500 test. Mutation data from solid tumors analyzed by clinical next-generation sequencing were abstracted from patients’ electronic medical records. Rows represent individual protein-level variants. Colored tiles indicate detection of a variant in the corresponding genomic source; gray tiles indicate that the variant was not detected. The bar plot at right indicates the number of genomic sources in which each variant was detected. The *ESR1* p.Y537S mutation (red font) in patient ME003 was detected in AI-TC-enriched cells, matched effusion cell-free DNA, and a liver metastasis collected within five days of the effusion, but was not detected in the primary tumor resected six years earlier (red box).

To assess genomic relatedness across tumor-derived analytes, variants detected by WES of AI-TC-enriched samples were compared with available genomic profiles from matched effusion cell-free DNA and solid tumors analyzed by UCSF500 or clinical next-generation sequencing (Fig. 5B). Multiple alterations detected in AI-TCs were also identified in independently profiled tumor-derived specimens from the same patients, demonstrating shared genomic features across analytes. Conversely, other alterations were detected in only a subset of matched specimens, revealing genomic divergence across tumor-derived analytes. For example, in patient ME003, the *ESR1* p.Y537S mutation was detected in AI-TC-enriched cells, matched effusion cell-free DNA, and a liver metastasis biopsy collected within five days of the effusion, but was not detected in the primary tumor resected six years earlier (Fig. 5B).

Together, these findings demonstrate that morphology-guided enrichment provides sufficient tumor-derived material for WES and enables mutation profiling and comparison with independently analyzed tumor-derived specimens.

### Longitudinal profiling reveals dynamic morphological and genomic heterogeneity in malignant effusions

To investigate temporal changes in tumor cell phenotype and genomic composition, we integrated longitudinal single-cell morphological and genomic profiling in patients with serial malignant effusion samples. Patients ME002 and ME014 were selected for detailed analysis because multiple serial samples were available together with matched genomic data from archival primary tumors and effusion cell-free DNA (Fig. 6). Longitudinal changes in AI-estimated TCF (Fig. S6A) and morphotype composition (Fig. S6B) were also observed in additional patients with serial malignant effusion samples.

**Figure 6.**
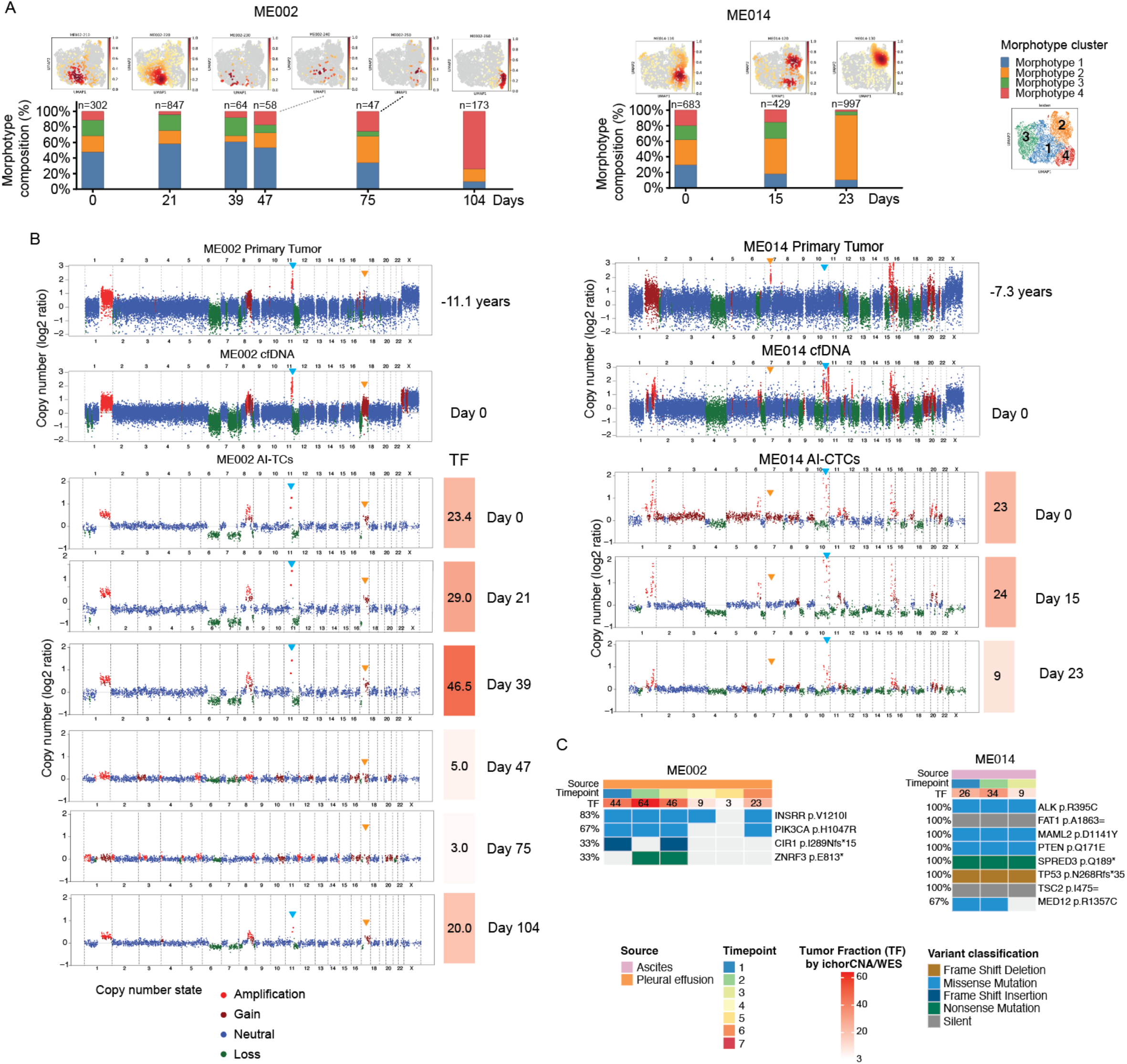
Longitudinal profiling reveals dynamic morphological and genomic heterogeneity in malignant effusions. **A.** Longitudinal changes in AI-identified tumor cell (AI-TC) morphotype composition in patients ME002 and ME014. Uniform manifold approximation and projection (UMAP) density plots are shown above corresponding stacked bars depicting the proportions of Morphotypes 1–4 at each collection timepoint. The number of AI-TCs analyzed at each timepoint is indicated above each bar. **B.** Longitudinal copy-number profiles of ME002 and ME014. Genome-wide copy-number profiles are shown for archival primary tumors obtained 11.1 and 7.3 years before malignant effusion collection for ME002 and ME014, respectively; matched malignant effusion cfDNA collected at day 0; and serial AI-TC-enriched samples. For ME002, AI-TC-enriched samples were collected on days 0, 21, 39, 47, 75, and 104; for ME014, samples were collected on days 0, 15, and 23. Copy-number states are indicated as amplification, gain, neutral, or loss. Tumor fractions (TFs) estimated by ichorCNA are indicated for each AI-TC-enriched sample. **C.** Longitudinal somatic mutation profiles detected by whole-exome sequencing (WES) of AI-TC-enriched samples from ME002 and ME014. Columns represent sequential samples and are annotated by fluid source, collection timepoint, and ichorCNA-estimated TF. Rows represent individual protein-level mutations in OncoKB cancer genes, and percentages on the left indicate the proportion of longitudinal samples from each patient in which each variant was detected. Tile color indicates variant type. Dark grey tiles are synonymous variants. Light grey tiles are undetected.

Patient ME002 had HR-positive/HER2-negative metastatic breast cancer and underwent repeated thoracentesis, during which six malignant pleural effusion samples were collected over 104 days. AI-powered single-cell analysis demonstrated marked changes in morphotype composition over time, with a progressive shift from predominantly Morphotype 1 toward increased representation of Morphotype 4 at later timepoints (Fig. 6A). Low-pass whole-genome sequencing of AI-TC-enriched samples identified multiple copy-number alterations, including focal amplification of 11q13 encompassing *CCND1* (Fig. 6B). Copy-number alteration signals were prominent at days 0, 21, and 39, markedly attenuated at days 47 and 75, and increased again at day 104, paralleling changes in sequencing-derived TF from 23.4%, 29.0%, and 46.5% at the first three timepoints to 5.0% and 3.0% at days 47 and 75, respectively, followed by an increase to 20.0% at day 104 (Fig. 6B).

Comparison with an archival primary tumor collected 11 years earlier revealed multiple shared copy-number alterations, including amplification of 11q13, supporting genomic relatedness between the primary tumor and effusion-derived tumor cells (Fig. 6B). However, additional alterations differed between specimens. In particular, a chromosome 17q gain detected in the AI-TCs was not evident in the archival primary tumor but was also detected in matched effusion cell-free DNA collected at day 0, demonstrating genomic divergence from the earlier primary tumor while providing independent support for the alteration in the metastatic effusion (Fig. 6B). Longitudinal WES further identified *PIK3CA* p.H1047R across four of six serial AI-TC-enriched samples, together with additional variants detected at subsets of timepoints (Fig. 6C, Fig. S6C).

Patient ME014 had HR-positive/HER2-negative metastatic breast cancer and underwent repeated paracentesis, during which three malignant ascites samples were collected over 23 days. Morphotype composition changed over this interval, with expansion of Morphotype 2 from day 0 to days 15 and 23 (Fig. 6A). Copy-number alteration profiling demonstrated numerous genomic alterations shared among the archival primary tumor collected 7.3 years earlier, effusion cell-free DNA, and serial AI-TC-enriched samples, consistent with genomic relatedness across tumor-derived specimens (Fig. 6B). Nevertheless, several alterations differed across specimens, including a chromosome 7 amplification apparent in the archival primary tumor but not in the effusion-derived samples, whereas focal amplifications on chromosome 10 were observed in the effusion-derived AI-TCs and cell-free DNA but not in the primary tumor (Fig. 6B). Longitudinal WES demonstrated persistent detection of several variants across all three effusion samples, including mutations in *ALK*, *PTEN*, and *TP53* (Fig. 6C, Fig. S6C).

Together, these longitudinal analyses demonstrate dynamic changes in tumor cell fraction and morphological composition while revealing both persistent and divergent genomic features across serial effusion samples and tumor-derived analytes collected years apart.

## DISCUSSION

Malignant effusions represent a clinically accessible source of metastatic tumor cells. However, background immune cells pose significant challenges for isolating and characterizing these cells. This study builds on prior work that established the feasibility of AI-powered, morphology-based enrichment of tumor cells from malignant effusions for downstream molecular characterization ^23^. Here, we extend this approach by integrating single-cell morphology information with morphotyping and genomic profiling to interrogate tumor identity and heterogeneity in these fluids.

Comparison of the AI-derived cell identities with standard flow cytometry showed moderate to strong correlations in the proportions of major immune cell populations, supporting the ability of morphology-based classification to distinguish major cell types within the complex cellular milieu of malignant effusions. In contrast, the proportions of AI-TCs were not concordant between the two methods. This discordance may be due in part to fundamental technical differences in sample preparation and in how tumor cells are defined for the two methodologies, as well as to potential misclassifications by either method. Whereas flow cytometry identified tumor cells using two epithelial cell-surface markers (EpCAM and CLDN4), the AI-based approach relied on cell morphology and did not require tumor cells to express a specific surface antigen. This distinction may matter in metastatic disease, where epithelial–mesenchymal plasticity can generate heterogeneous tumor cell states or reduce epithelial marker expression, potentially limiting marker-based detection of tumor cells ^41–43^.

Previous studies have demonstrated the feasibility of using tumor cells isolated directly from malignant effusions as a liquid-biopsy analyte for genomic profiling ^15,44^. Tiran et al. performed low-coverage whole-genome sequencing of cells isolated from malignant pleural effusions from patients with metastatic breast cancer, including phenotypically enriched tumor-cell subpopulations ^15^. Chen et al. subsequently used hexokinase 2 and cytokeratin-based identification to isolate tumor cells from malignant pleural effusions across multiple cancer types and performed single-cell DNA sequencing ^44^. However, both studies used fluorescently labeled markers to detect or enrich for tumor cells, including ALDH1 in Tiran et al. ^15^, and hexokinase 2 and cytokeratins in Chen et al. ^44^ In contrast, our label-free approach reduces reliance on predefined molecular or physical characteristics, potentially enabling representation of a wider spectrum of phenotypically heterogeneous tumor cells for molecular profiling.

Our analysis identified four tumor cell morphotypes, a classification based solely on image-derived phenotypic clusters rather than on discrete molecular or functional cell states. We observed heterogeneity in morphotype composition across patients and in serial samples from the same patient. Longitudinal changes in morphotype composition within individual patients suggest that tumor cell morphology may be a dynamic biomarker that could be monitored over time. Nonetheless, whether these morphotypes reflect distinct tumor cell biology warrants further investigation.

Morphology-guided, label-free enrichment and cell sorting enabled genomic characterization of AI-TC-enriched samples. Copy-number and mutation analyses identified genomic alterations that provided orthogonal genomic support for the tumor origin of AI-TC-enriched populations. Comparison with genomic data from matched analytes (including cell-free DNA from the same effusion fluids and solid tumors from the same patient) demonstrated both genomic relatedness and divergence. Importantly, longitudinal analysis showed that sequencing-derived TF influenced the strength of the genomic signal. Samples with higher TF generally exhibited more readily detectable copy-number alterations, whereas samples with low TF showed attenuated genomic signals and, in some cases, fell below the detection limits of sequencing-based approaches. These observations highlight a fundamental challenge in genomic profiling of malignant effusions: highly abundant nonmalignant cells can dilute tumor-derived DNA. Increasing sequencing-derived TF through morphology-guided enrichment may therefore improve the recovery of tumor-associated genomic information, particularly from specimens with substantial nonmalignant-cell admixture.

These findings further support malignant effusions as a source of cellular and cell-free analytes for liquid biopsy of metastatic disease. Because malignant effusions can be repeatedly collected during therapeutic drainage, they provide an opportunity to characterize the contemporary genomic landscape of metastatic disease without requiring an additional invasive procedure. This potential is illustrated by patient ME003, in whom *ESR1* p.Y537S was detected in AI-TC-enriched cells, matched effusion cell-free DNA, and a contemporaneous liver metastasis, but not in the primary tumor resected six years earlier. *ESR1* ligand-binding-domain mutations are frequently selected during endocrine therapy and represent an established mechanism of acquired resistance to aromatase inhibitors ^45^. Similarly, *ERBB2* amplification was detected in both AI-TC-enriched cells and effusion cell-free DNA from patient ME006, whose primary tumor had been clinically classified as HER2-negative. Together, these examples illustrate how genomic profiling of malignant effusions may capture clinically relevant features of contemporary metastatic disease that are not represented by historical tumor characterization and could potentially inform treatment selection. Whether longitudinal changes in TCF or morphotype composition reflect treatment response, clonal selection, or clinical outcome will require evaluation in prospectively collected cohorts with detailed treatment and outcome data.

### Limitations

The study has several limitations. The cohort was small and heterogeneous, and included only patients with metastatic breast cancer who received different treatments. The availability of serial malignant effusion samples, matched cell-free DNA and solid tumors for genomic comparisons varied. Because sequencing was performed on whole-genome-amplified DNA, amplification-related coverage nonuniformity, allelic dropout, and sequencing artifacts may have contributed to apparent differences in copy number and variant detection across serial samples. To confirm alterations detected in AI-TC-enriched fractions, we used genomic data from cell-free DNA isolated from the same fluid and matched solid tumors, which were not subjected to whole-genome amplification. Nonetheless, longitudinal differences in individual genomic alterations should not necessarily be interpreted as evidence of clonal evolution without independent validation. The sensitivity of genomic profiling was limited in samples with low sequencing-derived TF, and the absence of detectable copy-number alterations cannot distinguish insufficient tumor content from copy-number-neutral tumor genomes or other technical limitations of low-pass sequencing. Independent validation of the morphology-based classifier in a larger cohort and against additional orthogonal measures of tumor cell identity will be important. Whether morphotypes represent discrete tumor biological states remains unknown. Future work includes single-cell RNA sequencing of AI-TC-enriched samples to link individual morphotypes with genomic and transcriptomic states.

## Conclusions

Despite the potential value of malignant effusions as a source of metastatic tumor material, these fluids remain an underutilized biospecimen. Characterizing tumor cells in malignant effusions can provide longitudinal snapshots of metastatic disease and help identify novel biomarkers and therapeutic targets for advanced cancers. Our approach converts routinely obtained, often discarded malignant-effusion material into a multidimensional source of information on metastatic tumor heterogeneity. The AI-powered single-cell morphology analysis platform enabled label-free identification, phenotyping, and enrichment of putative tumor cells, and genomic analysis provided orthogonal evidence for tumor origin of enriched populations while revealing phenotypic and genomic heterogeneity over time.

## DECLARATIONS

### Funding

This work was supported by the California Breast Cancer Research Program (CBCRP) IDEA Award (M.J.M.) and the National Cancer Institute of the National Institutes of Health (R01CA292019; M.J.M., J.R., and M.M.). Deepcell Inc. provided technical and sequencing support at no cost.

## Author Contributions

Conceptualization: M.J.M.M., L.A.H., M.R., C.J., M.M., R.B., H.S.R., L.v.V., R.W.S., M.S., M.Mas., J.R., H.G.; Methodology: M.J.M.M., J.Cr., J.G., S.Z., K.Y., P.S., N.G., L.Hs., M.W., M.P., C.J., J.N., E.C.S., A.M., A.P., J.Ca., M.B., L.He., V.B., A.M.C.; Software: R.C.; Formal analysis: M.J.M.M., J.Cr., N.G., R.C., J.K., Z.L., S.M., R.W.S.; Resources: M.J.M.M., L.A.H., M.M., H.S.R., L.v.V., P.D., M.S., M.Mas., J.R., H.G.; Data curation: N.G., R.C., J.K., Z.L.; Writing – original draft: M.J.M.M., J.R.; Writing – review & editing: M.J.M.M., N.G., A.L.D., A.J., R.W.S.; Visualization: M.J.M.M., N.G., R.C., Z.L., S.M., J.R.; Supervision: M.J.M.M., A.M., A.J., J.R., H.G.; Project administration: M.J.M.M., L.A.H., M.R., C.J., M.S., M.Mas., J.R., H.G.; Funding acquisition: M.J.M.M., L.A.H., J.R.; Patient advocacy: A.L.D., C.S. All authors reviewed and approved the final manuscript.

## Data Availability

Genomic sequencing data will be deposited in an appropriate controlled-access repository, subject to institutional approvals and patient consent. Additional de-identified data supporting the findings of this study are available from the corresponding authors upon reasonable request, subject to institutional and ethical restrictions.

## Acknowledgments

We thank Colin Chandler for computational assistance and Lauryn Horita for assistance with patient-related research activities. This work is dedicated to the memory of Carol Simmons, in recognition of her contributions to breast cancer research and advocacy.

## Competing interests

J. Cr., R.C., J.K., L.Hs., M.W., Z.L., S.Man., M.P., M.R., C.Jo., J.N., E.C.S., A.M., A.J., M.Sal., and M.Mas. are current or former employees of Deepcell Inc.

## Code availability

No custom code was developed for this study. All computational analyses were performed using existing software and publicly available tools, as described in the Methods.

## Ethics statement

This study was conducted in accordance with protocols approved by the University of California, San Francisco Institutional Review Board (IRB). Details regarding ethical approval and informed consent are provided in the Methods section.

## Notes

### Author Declarations

The University of California, San Francisco Institutional Review Board (IRB) gave ethical approval for this work.

