## Supplementary Information for "AI-powered single-cell imaging and morphology-guided label-free enrichment reveal tumor heterogeneity, genomic relatedness, and divergence in malignant effusions"

### SUPPLEMENTARY FIGURES

A

B

C

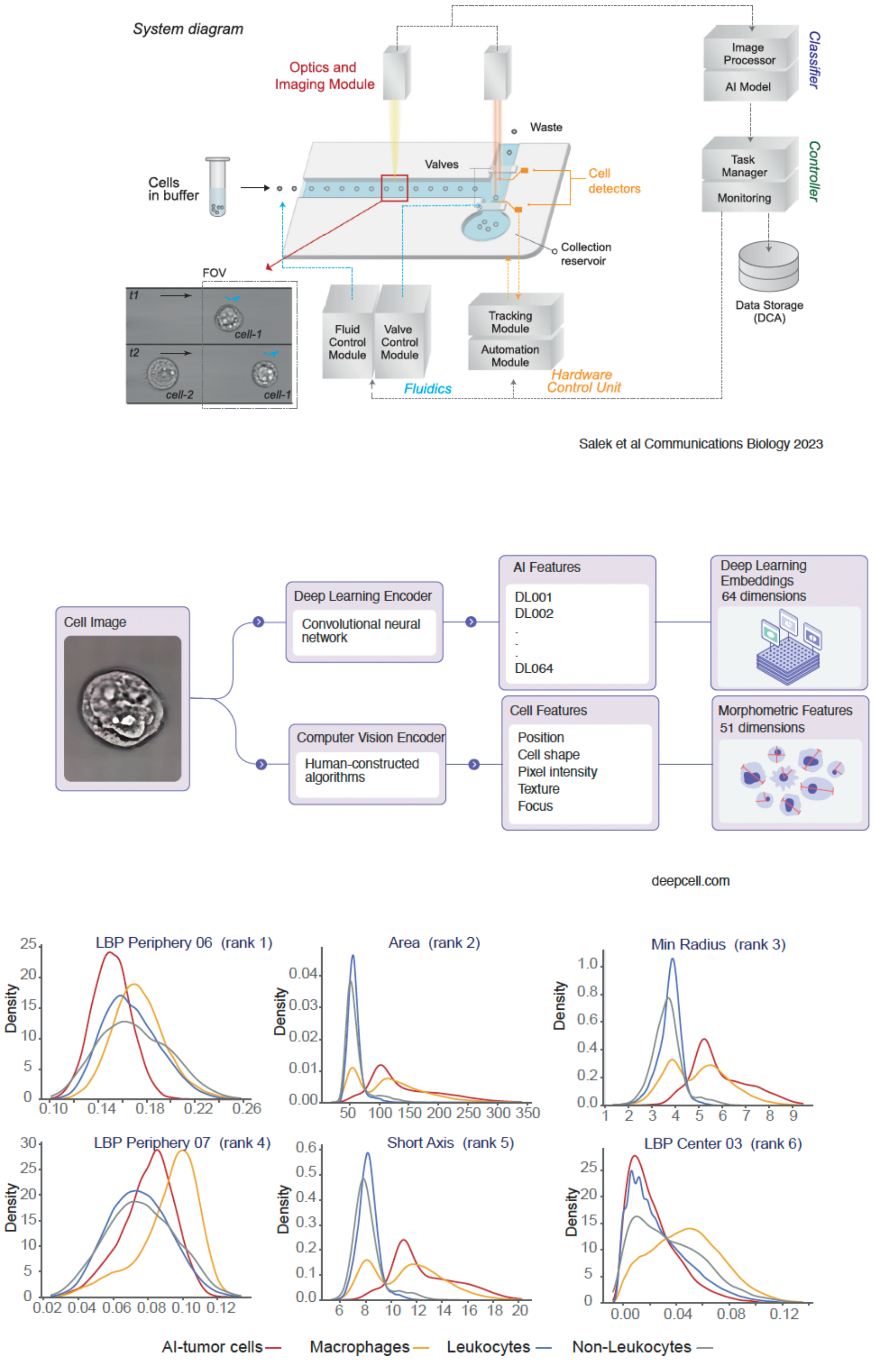


**Supplementary Figure 1. Imaging platform and image-derived cellular features.**

**A.** Schematic of the Deepcell platform illustrating cell imaging, real-time cell detection and tracking, AI-powered classification, and fluidic sorting. Adapted from Salek et al. 2023.

**B.** Schematic illustrating extraction of image-derived features from individual cell images. A deep-learning encoder generates 64-dimensional deep-learning embeddings, whereas computer-vision algorithms generate 51 morphometric features describing cell position, shape, pixel intensity, texture, and focus.

**C.** Density distributions of the six highest-ranked morphometric features distinguishing AI-identified tumor cells, macrophages, leukocytes (non-macrophage), and non-leukocytes: local binary pattern (LBP) Periphery 06, area, minimum radius, LBP Periphery 07, short axis, and LBP Center 03. See Table S4 for definitions of morphometric features.

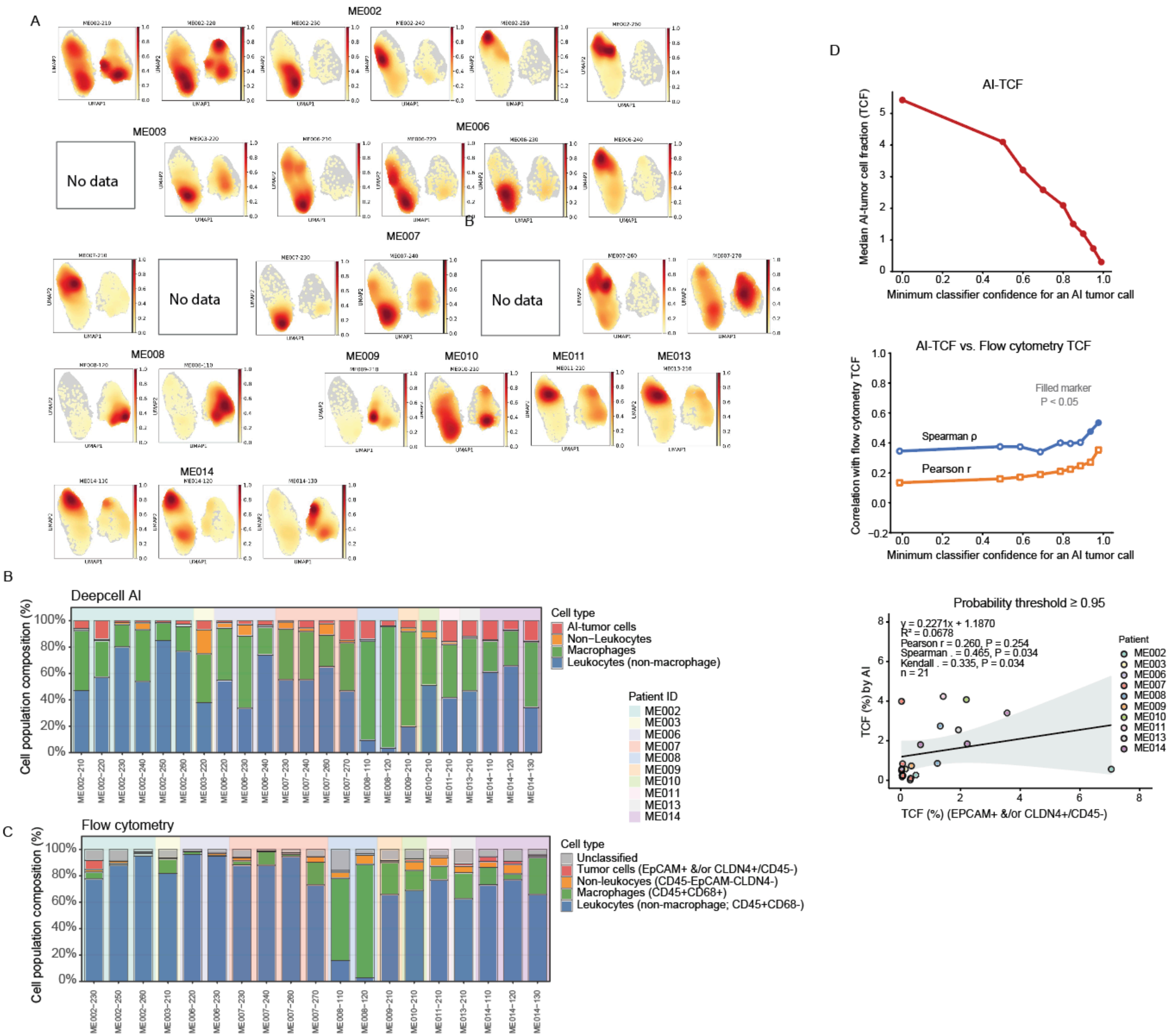
A
ME002
ME003
No data
ME006
ME007
No data
No data
ME008
ME009
ME010
ME011
ME013
ME014
UMAP1
UMAP2
B
Deepcell AI
Cell population composition (%)
100%
80%
60%
40%
20%
0%
Cell type
AI-tumor cells
Non−Leukocytes
Macrophages
Leukocytes (non-macrophage)
Patient ID
ME002
ME003
ME006
ME007
ME008
ME009
ME010
ME011
ME013
ME014
ME002−210
ME002−220
ME002−230
ME002−240
ME002−250
ME002−260
ME003−220
ME006−220
ME006−230
ME006−240
ME007−230
ME007−240
ME007−260
ME007−270
ME008−110
ME008−120
ME009−210
ME010−210
ME011−210
ME013−210
ME014−110
ME014−120
ME014−130
C
Flow cytometry
Cell population composition (%)
Cell type
Unclassified
Tumor cells (EpCAM+ &/or CLDN4+/CD45-)
Non-leukocyes (CD45-EpCAM-CLDN4-)
Macrophages (CD45+CD68+)
Leukocytes (non-macrophage; CD45+CD68-)
ME002−230
ME002−250
ME002−260
ME003−210
ME006−220
ME006−230
ME007−230
ME007−240
ME007−260
ME007−270
ME008−110
ME008−120
ME009−210
ME010−210
ME011−210
ME013−210
ME014−110
ME014−120
ME014−130
D
AI-TCF
Median AI-tumor cell fraction (TCF)
Minimum classifier confidence for an AI tumor call
AI-TCF vs. Flow cytometry TCF
Correlation with flow cytometry TCF
Filled marker
P < 0.05
Spearman ρ
Pearson r
Minimum classifier confidence for an AI tumor call
Probability threshold ≥ 0.95
y = 0.2271x + 1.1870
R² = 0.0678
Pearson r = 0.260, P = 0.254
Spearman . = 0.465, P = 0.034
Kendall . = 0.335, P = 0.034
n = 21
TCF (%) by AI
TCF (%) (EPCAM+ &/or CLDN4+/CD45-)
Patient
ME002
ME003
ME006
ME007
ME008
ME009
ME010
ME011
ME013
ME014

**Supplementary Figure 2. Sample-level characterization of malignant effusion cell populations by AI and flow cytometry.**

**A.** Density distributions of cells in uniform manifold approximation and projection (UMAP) space for individual malignant effusion samples, grouped by patient. Samples without available data are indicated.

**B.** Cell-population composition of individual malignant effusion samples determined by AI. Stacked bars show the proportions of leukocytes (non-macrophage), macrophages, non-leukocytes, and AI-identified tumor cells.

**C.** Cell-population composition of individual samples determined by flow cytometry. Stacked bars show leukocytes (non-macrophage; CD45+/CD68-), macrophages (CD45+/CD68+), tumor cells (EpCAM+ and/or CLDN4+/CD45-), non-leukocytes (CD45-/EpCAM-/CLDN4-), and unclassified cells. Background shading identifies patients.

**D.** Effect of AI classifier confidence thresholds on tumor-cell fraction (TCF) estimates and concordance with flow cytometry. In a post hoc exploratory analysis, we evaluated whether applying increasingly stringent minimum classifier confidence thresholds for AI-based tumor-cell identification improved concordance with flow cytometry-derived TCF estimates, compared with classification based on the highest predicted probability. Top: Median AI-derived TCF decreased progressively with increasing classifier confidence thresholds. Middle: Pearson and Spearman correlations between AI-derived and flow cytometry-derived TCF generally increased at higher confidence thresholds, with statistically significant correlations observed at the most stringent thresholds (filled markers, $P < 0.05$). Bottom: Scatter plot comparing AI-derived and flow cytometry-derived TCF at a minimum classifier confidence threshold of 0.95 ($n = 21$ samples). Points represent individual samples colored by patient. The solid line represents linear regression, and the shaded region indicates the 95% confidence interval. Pearson, Spearman, and Kendall correlation coefficients, corresponding P values, the regression equation, and $R^2$ are shown.

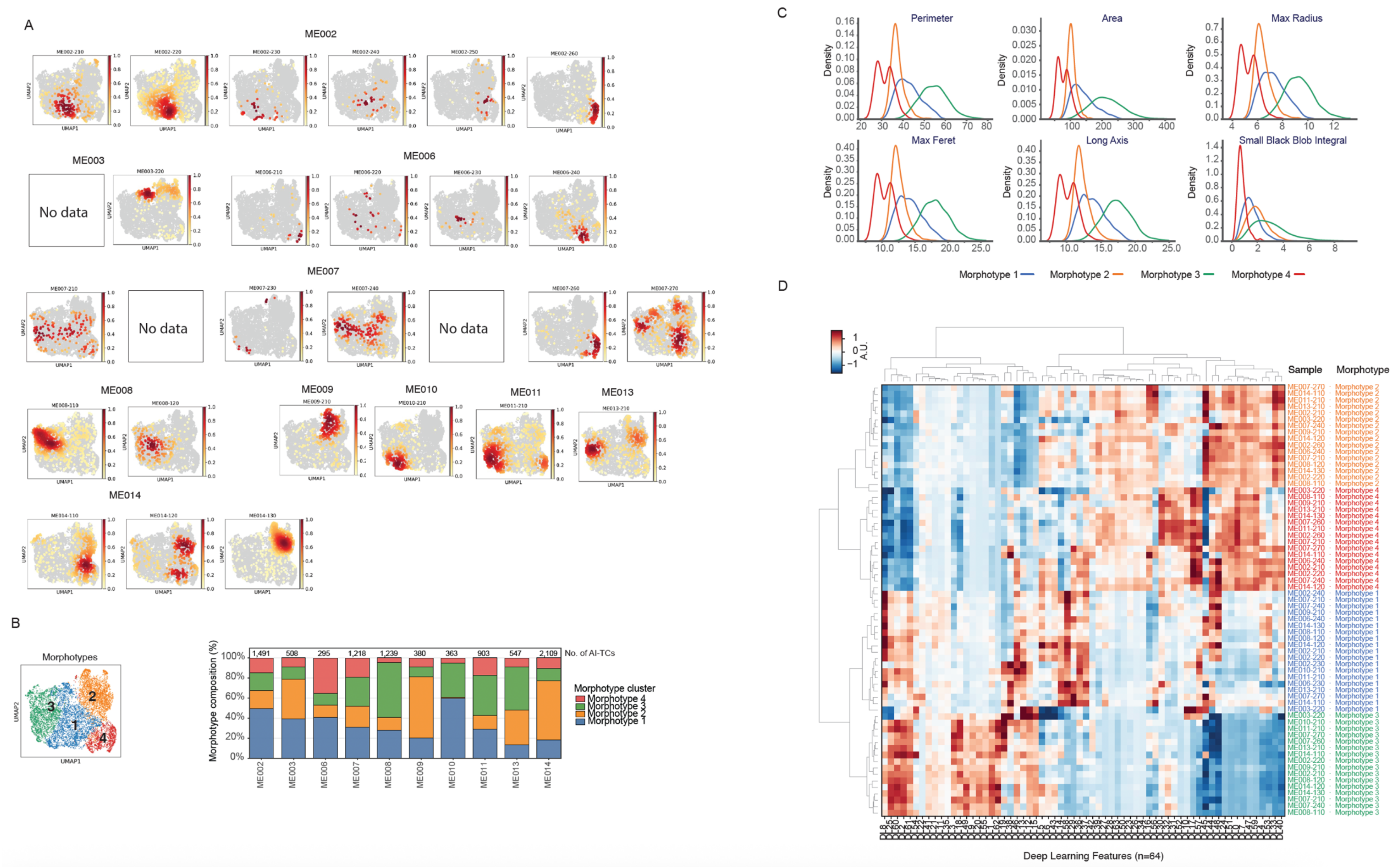
A
ME002
ME003
No data
ME006
ME007
No data
No data
ME008
ME009
ME010
ME011
ME013
ME014
UMAP1
UMAP2
B
Morphotypes
1
2
3
4
Morphotype composition (%)
No. of AI-TCs
1,491
508
295
1,218
1,239
380
363
903
547
2,109
ME002
ME003
ME006
ME007
ME008
ME009
ME010
ME011
ME013
ME014
Morphotype cluster
Morphotype 4
Morphotype 3
Morphotype 2
Morphotype 1
C
Perimeter
Area
Max Radius
Max Feret
Long Axis
Small Black Blob Integral
Density
Morphotype 1
Morphotype 2
Morphotype 3
Morphotype 4
D
A.U.
Sample
Morphotype
Deep Learning Features (n=64)

**Supplementary Figure 3. Distribution and deep-learning characteristics of AI-identified tumor cell morphotypes across malignant effusion samples.**

**A.** Sample-specific density distributions of AI-identified tumor cells in uniform manifold approximation and projection (UMAP) space, grouped by patient. Samples without available data are indicated.

**B.** Morphotype composition of AI-TCs for each patient. Stacked bars show the proportions of Morphotypes 1–4, and the total number of AI-TCs analyzed for each patient is indicated above each bar. For patients with serial samples, AI-TCs were aggregated AI-TCs across all available timepoints.

**C.** Density distributions of selected morphometric features across the four morphotype clusters, including perimeter, area, maximum radius, maximum Feret diameter, long-axis length, and small-black-blob integral.

**D.** Hierarchically clustered heatmap of deep-learning image features for each sample-by-morphotype combination. Rows represent morphotype clusters within individual samples and columns represent the 64 deep-learning features; values are standardized feature measurements shown in arbitrary units.

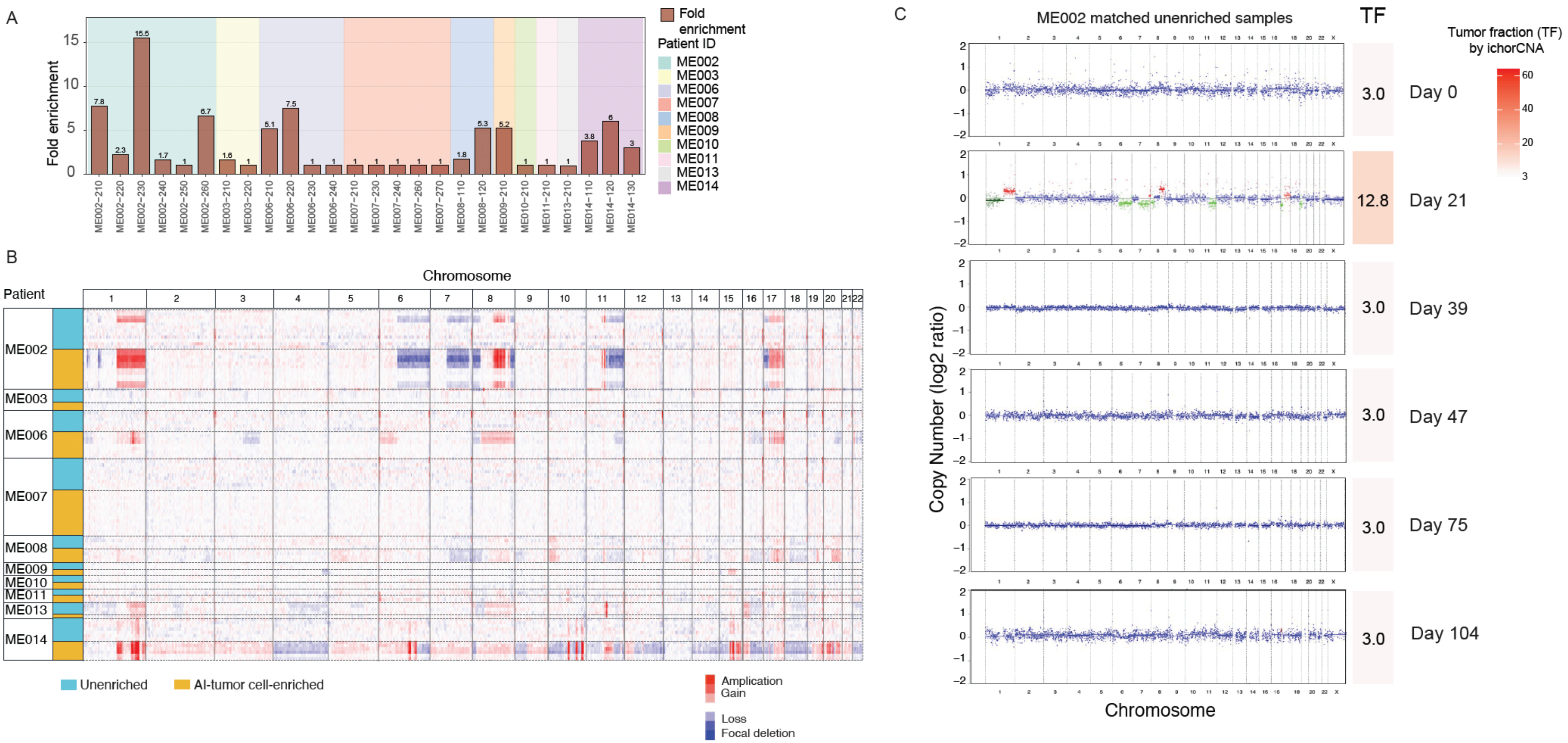

A
Fold enrichment
Fold enrichment
Patient ID
ME002
ME003
ME006
ME007
ME008
ME009
ME010
ME011
ME013
ME014
ME002-210
ME002-220
ME002-230
ME002-240
ME002-250
ME002-260
ME003-210
ME003-220
ME006-210
ME006-220
ME006-230
ME006-240
ME007-210
ME007-230
ME007-240
ME007-260
ME007-270
ME008-110
ME008-120
ME009-210
ME010-210
ME011-210
ME013-210
ME014-110
ME014-120
ME014-130
7.8
2.3
15.5
1.7
1
6.7
1.6
1
5.1
7.5
1
1
1
1
1
1
1
1.8
5.3
5.2
1
1
1
3.8
6
3
B
Chromosome
Patient
ME002
ME003
ME006
ME007
ME008
ME009
ME010
ME011
ME013
ME014
Unenriched
AI-tumor cell-enriched
Amplication
Gain
Loss
Focal deletion
C
ME002 matched unenriched samples
TF
Tumor fraction (TF) by ichorCNA
3.0
Day 0
12.8
Day 21
3.0
Day 39
3.0
Day 47
3.0
Day 75
3.0
Day 104
Copy Number (log2 ratio)
Chromosome

**Supplementary Figure 4. Copy-number profiles and tumor enrichment following morphology-guided isolation of AI-identified tumor cells (AI-TC).**

**A.** Fold enrichment of tumor fraction (TF) following morphology-guided AI-TC enrichment. Fold enrichment was calculated as the ratio of ichorCNA-estimated TF in the AI-TC-enriched fraction to that in the corresponding unenriched sample; values are shown above individual bars. Samples with TF below the 3% lower limit of detection [31] were assigned 3% for this calculation.

**B.** Genome-wide heatmap of copy-number alterations detected by low-pass whole-genome sequencing in paired unenriched and AI-TC-enriched samples. Samples are grouped by patient, and copy-number states are indicated as amplification, gain, loss, and focal deletion.

**C.** Genome-wide copy-number profiles of six serial unenriched malignant pleural effusion samples from patient ME002 collected over 104 days. TF estimated by ichorCNA is shown for each sample. Genome-wide copy-number profiles of matched AI-TC-enriched samples for ME002 are shown in Figure 6.

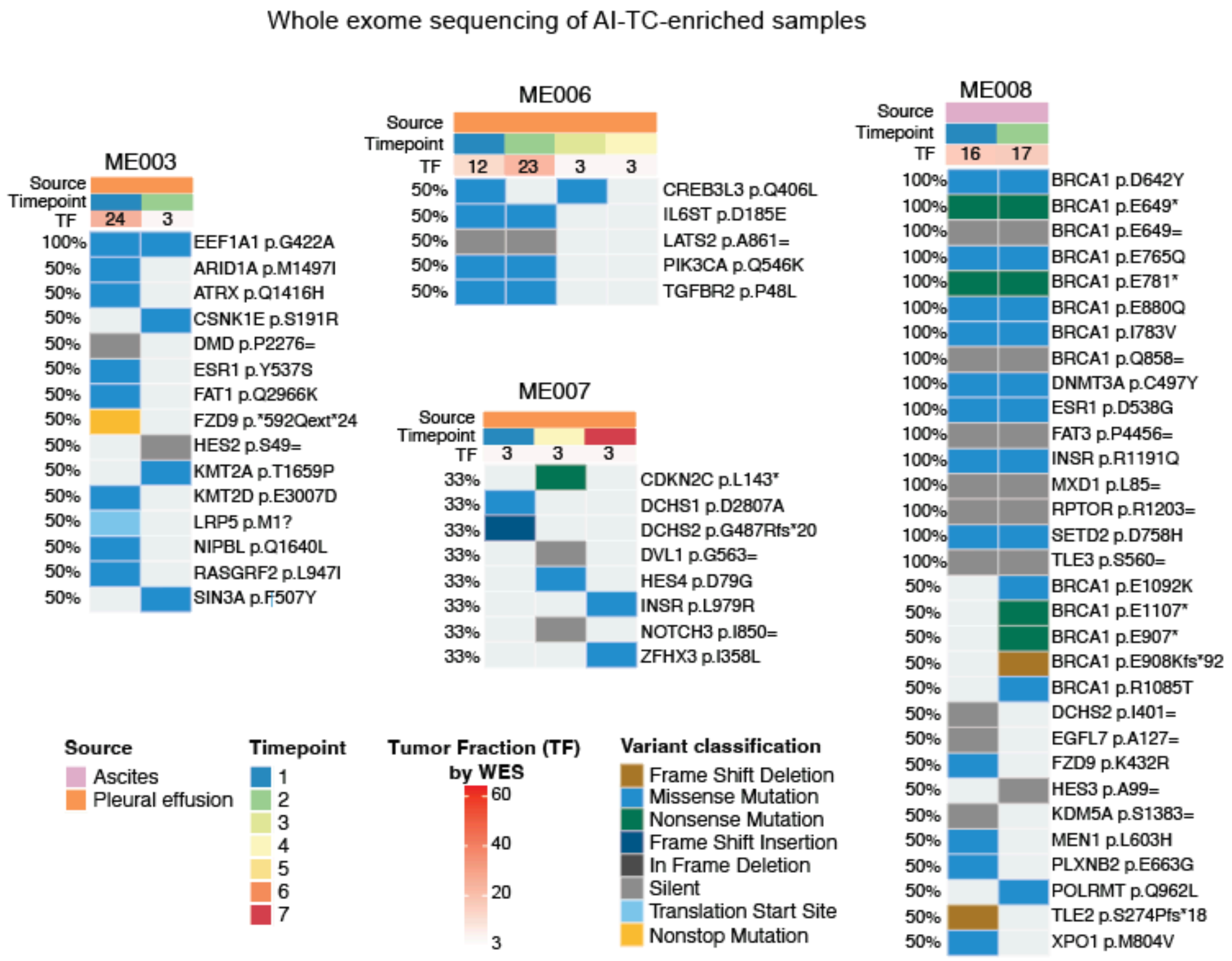


**Supplementary Figure 5. Longitudinal mutation profiles in patients with serial malignant effusion samples.**

Longitudinal somatic mutation profiles detected by whole-exome sequencing (WES) of AI-tumor cell (AI-TC)-enriched samples from patients with serial malignant effusion collections. Results for ME002 and ME014 are shown in Figure 6. Columns represent sequential samples and are annotated by fluid source, collection timepoint, and ichorCNA-estimated tumor fraction (TF). Rows represent individual protein-level mutations in OncoKB cancer genes, and the percentage on the left indicates the proportion of longitudinal samples from each patient in which the variant was detected. Tile color indicates variant type. Dark grey tiles are synonymous variants. Light grey tiles are undetecte

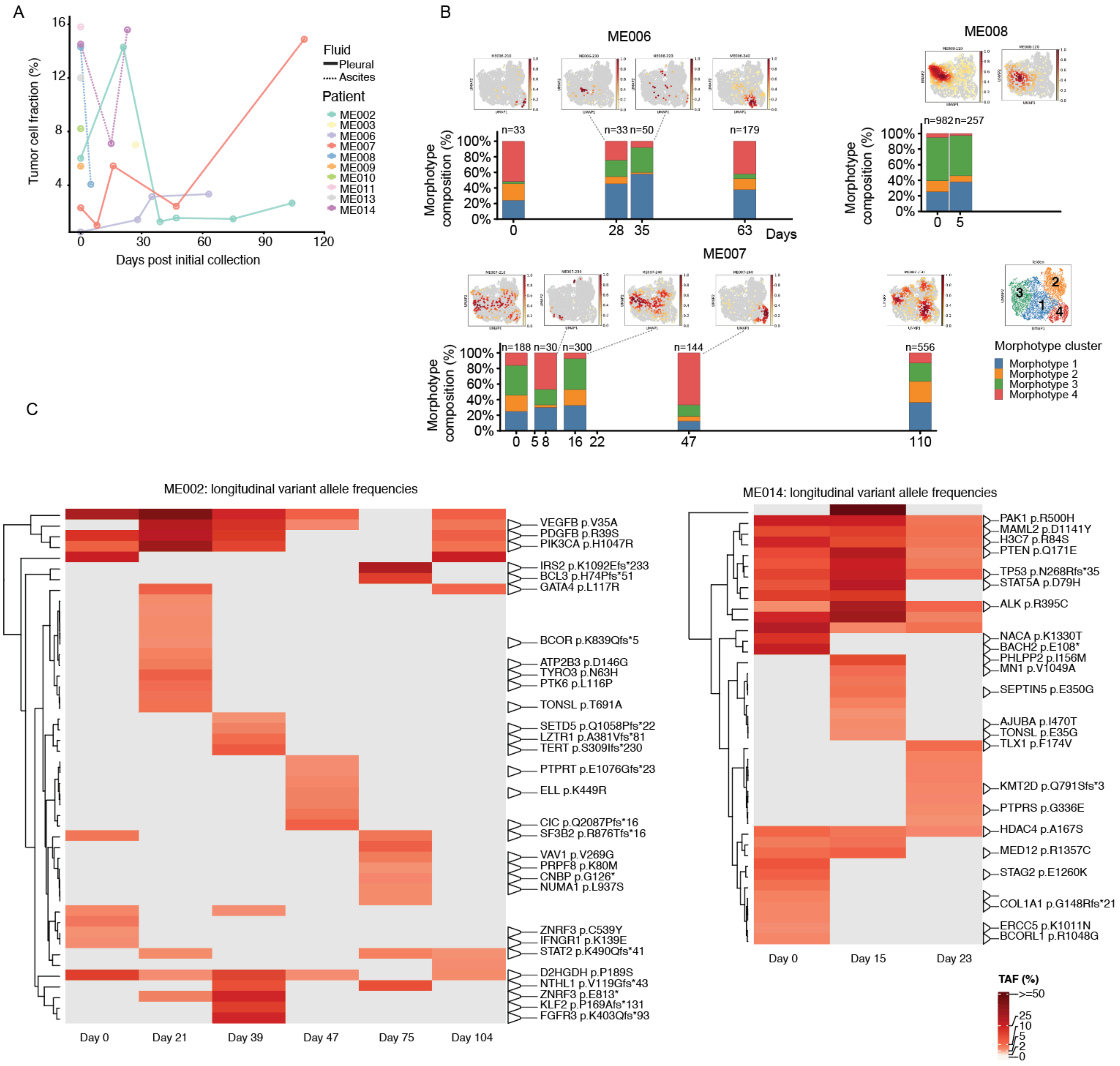
A
Tumor cell fraction (%)
Days post initial collection
Fluid
Pleural
Ascites
Patient
ME002
ME003
ME006
ME007
ME008
ME009
ME010
ME011
ME013
ME014
B
ME006
ME008
ME007
Morphotype composition (%)
Days
n=33
n=33
n=50
n=179
n=982
n=257
n=188
n=30
n=300
n=144
n=556
Morphotype cluster
Morphotype 1
Morphotype 2
Morphotype 3
Morphotype 4
C
ME002: longitudinal variant allele frequencies
VEGFB p.V35A
PDGFB p.R39S
PIK3CA p.H1047R
IRS2 p.K1092Efs*233
BCL3 p.H74Pfs*51
GATA4 p.L117R
BCOR p.K839Qfs*5
ATP2B3 p.D146G
TYRO3 p.N63H
PTK6 p.L116P
TONSL p.T691A
SETD5 p.Q1058Pfs*22
LZTR1 p.A381Vfs*81
TERT p.S309Ifs*230
PTPRT p.E1076Gfs*23
ELL p.K449R
CIC p.Q2087Pfs*16
SF3B2 p.R876Tfs*16
VAV1 p.V269G
PRPF8 p.K80M
CNBP p.G126*
NUMA1 p.L937S
ZNRF3 p.C539Y
IFNGR1 p.K139E
STAT2 p.K490Qfs*41
D2HGDH p.P189S
NTHL1 p.V119Gfs*43
ZNRF3 p.E813*
KLF2 p.P169Afs*131
FGFR3 p.K403Qfs*93
Day 0
Day 21
Day 39
Day 47
Day 75
Day 104
ME014: longitudinal variant allele frequencies
PAK1 p.R500H
MAML2 p.D1141Y
H3C7 p.R84S
PTEN p.Q171E
TP53 p.N268Rfs*35
STAT5A p.D79H
ALK p.R395C
NACA p.K1330T
BACH2 p.E108*
PHLPP2 p.I156M
MN1 p.V1049A
SEPTIN5 p.E350G
AJUBA p.I470T
TONSL p.E35G
TLX1 p.F174V
KMT2D p.Q791Sfs*3
PTPRS p.G336E
HDAC4 p.A167S
MED12 p.R1357C
STAG2 p.E1260K
COL1A1 p.G148Rfs*21
ERCC5 p.K1011N
BCORL1 p.R1048G
Day 0
Day 15
Day 23
TAF (%)
>=50

**Supplementary Figure 6. Longitudinal changes in AI-identified tumor cell fraction and morphotype composition.**

**A.** Longitudinal tumor cell fraction of AI-identified tumor cells (AI-TC) in unenriched samples across all patients.

**B.** Longitudinal changes in AI-TC morphotype composition in patients ME006, ME007, and ME008. Plots for ME002 and ME014 are shown in Figure 6. Uniform manifold approximation and projection (UMAP) density plots are shown above corresponding stacked bars depicting the proportions of morphotype clusters 1–4 at each collection timepoint. The number of AI-identified tumor cells analyzed (*n*) is indicated above each bar, and the x-axis indicates days from the initial sample collection.

**C.** Longitudinal genomic alterations in serial malignant effusion samples. Heatmaps show longitudinal tumor allele frequencies (TAFs) of mutations in OncoKB cancer genes for individual patients. Rows represent individual genomic alterations and columns represent serial malignant effusion samples ordered chronologically by collection day. Cell color indicates TAF, with increasing red intensity representing higher TAF; grey indicates that the alteration was not detected (ND) at that timepoint. Rows were ordered by unsupervised hierarchical clustering based on TAF profiles across serial samples, whereas columns were not clustered. Selected nonsilent alterations are annotated with the gene symbol and protein-level amino acid change, when available. Solid lines connect annotations to their corresponding alteration rows. Silent and other variant classes are included in the heatmaps but are not annotated. The same TAF color scale is applied across all patients.

#### Supplementary Tables

Table 1: Patient and tumor characteristics

| Characteristic | | n | % |
|---|---|---|---|
| **Subtype** | HR-positive/HER2-negative | 7 | 70% |
| | TNBC | 3 | 30% |
| | HER2-positive | 0 | 0% |
| **Effusion type** | Pleural | 8 | 80% |
| | Ascites | 2 | 20% |
| **Race** | White | 6 | 60% |
| | Asian | 3 | 30% |
| | Other | 1 | 10% |
| **Age** | Mean (range), years | 61 (47-80) | |

*Abbreviations:*
HR, hormone receptor; HER2, human epidermal growth factor receptor 2; TNBC, triple-negative breast cancer.

### Table 2: Samples and assays

| Patient ID | Sample ID | Effusion type | Timepoint | Collection interval (days) | Days from initial sample | Flow cytometry | Imaging | No. of cells inpur for whole genome amplification | Low-pass whole-genome sequencing | Whole-exome sequencing |
|---|---|---|---|---|---|---|---|---|---|---|
| ME002 | ME002-210 | Pleural | 1 | 0 | 0 | n.a. | • | 265 | • | • |
| | ME002-220 | Pleural | 2 | 21 | 21 | n.a. | • | 325 | • | • |
| | ME002-230 | Pleural | 3 | 18 | 39 | • | • | 217 | • | • |
| | ME002-240 | Pleural | 4 | 8 | 47 | n.a. | • | 250 | • | • |
| | ME002-250 | Pleural | 5 | 28 | 75 | • | • | 300 | • | • |
| | ME002-260 | Pleural | 6 | 29 | 104 | • | • | 250 | • | • |
| ME003 | ME003-210 | Pleural | 1 | 0 | 0 | n.a. | n.a. | 500 | • | • |
| | ME003-220 | Pleural | 2 | 27 | 27 | • | • | 250 | • | • |
| ME006 | ME006-210 | Pleural | 1 | 0 | 0 | • | • | 300 | • | • |
| | ME006-220 | Pleural | 2 | 28 | 28 | • | • | 350 | • | • |
| | ME006-230 | Pleural | 3 | 7 | 35 | • | • | 240 | • | • |
| | ME006-240 | Pleural | 4 | 28 | 63 | n.a. | • | 350 | • | • |
| ME007 | ME007-210 | Pleural | 1 | 0 | 0 | • | • | 260 | • | • |
| | ME007-220 | Pleural | 2 | 5 | 5 | n.a. | n.a. | n.a. | n.a. | n.a. |
| | ME007-230 | Pleural | 3 | 3 | 8 | • | • | 200 | • | n.a. |
| | ME007-240 | Pleural | 4 | 8 | 16 | • | • | 250 | • | • |
| | ME007-250 | Pleural | 5 | 6 | 22 | n.a. | n.a. | n.a. | n.a. | n.a. |
| | ME007-260 | Pleural | 6 | 25 | 47 | • | • | 300 | • | n.a. |
| | ME007-270 | Pleural | 7 | 63 | 110 | • | • | n.a. | • | • |
| ME008 | ME008-110 | Ascites | 1 | 0 | 0 | • | • | 290 | • | • |
| | ME008-120 | Ascites | 2 | 5 | 5 | • | • | 290 | • | • |
| ME009 | ME009-210 | Pleural | 1 | n.a. | n.a. | • | • | 250 | • | • |
| ME010 | ME010-210 | Pleural | 1 | n.a. | n.a. | • | • | 235 | • | • |
| ME011 | ME011-210 | Pleural | 1 | n.a. | n.a. | • | • | 250 | • | • |
| ME013 | ME013-210 | Pleural | 1 | n.a. | n.a. | • | • | 250 | • | • |
| ME014 | ME014-110 | Ascites | 1 | 0 | 0 | • | • | 300 | • | • |
| | ME014-120 | Ascites | 2 | 15 | 15 | • | • | 300 | • | • |
| | ME014-130 | Ascites | 3 | 8 | 23 | • | • | 350 | • | • |
| | | | | | **Total** | **21** | **25** | | **26** | **24** |

Table 3: Morphometric and deep-learning features associated with AI-tumor cell morphotypes

| Morphotype 1 | | Morphotype 2 | | Morphotype 3 | | Morphotype 4 | |
|---|---|---|---|---|---|---|---|
| **Feature** | **Importance** | **Feature** | **Importance** | **Feature** | **Importance** | **Feature** | **Importance** |
| Deep Learning Feature 58 | 6.94 | Deep Learning Feature 63 | 6.23 | MAX RADIUS | 9.26 | SMALL BLACK BLOB INTEGRAL | 7.40 |
| Deep Learning Feature 61 | 4.79 | Deep Learning Feature 33 | 5.74 | MAX FERET | 7.19 | MAX RADIUS | 6.60 |
| HU01 | 4.73 | Deep Learning Feature 12 | 5.25 | Deep Learning Feature 51 | 6.33 | MAX FERET | 5.92 |
| STANDARD DEVIATION | 2.80 | Deep Learning Feature 7 | 4.78 | PERIMETER | 6.01 | PERIMETER | 5.20 |
| MAX RADIUS | 2.55 | Deep Learning Feature 60 | 4.00 | LONG AXIS | 4.62 | Deep Learning Feature 61 | 5.04 |
| Deep Learning Feature 37 | 2.54 | Deep Learning Feature 46 | 3.93 | Deep Learning Feature 33 | 4.01 | LONG AXIS | 4.62 |
| Deep Learning Feature 13 | 2.21 | Deep Learning Feature 59 | 3.82 | AREA | 3.90 | LBP PERIPHERY 07 | 4.44 |
| Deep Learning Feature 36 | 2.21 | HU01 | 3.58 | Deep Learning Feature 0 | 3.43 | AREA | 4.08 |
| PERIMETER | 2.01 | Deep Learning Feature 1 | 3.07 | MIN FERET | 2.97 | Deep Learning Feature 0 | 3.85 |
| Deep Learning Feature 9 | 1.82 | STANDARD DEVIATION | 2.90 | SHORT AXIS | 2.91 | Deep Learning Feature 17 | 2.42 |
| Deep Learning Feature 8 | 1.80 | MIN FERET | 2.63 | Deep Learning Feature 59 | 2.25 | LBP PERIPHERY 03 | 2.36 |
| MAX FERET | 1.78 | Deep Learning Feature 24 | 2.45 | Deep Learning Feature 48 | 2.16 | MIN FERET | 2.34 |
| Deep Learning Feature 33 | 1.67 | SMALL BLACK BLOB INTEGRAL | 2.24 | Deep Learning Feature 7 | 2.01 | Deep Learning Feature 2 | 2.12 |
| MIN RADIUS | 1.59 | SHORT AXIS | 2.18 | Deep Learning Feature 2 | 1.84 | SHORT AXIS | 2.11 |
| SMALL BLACK BLOB INTEGRAL | 1.58 | Deep Learning Feature 0 | 2.14 | Deep Learning Feature 24 | 1.67 | LBP PERIPHERY 09 | 1.93 |
| LONG AXIS | 1.57 | MIN RADIUS | 2.01 | MIN RADIUS | 1.46 | HU01 | 1.69 |
| SHORT AXIS | 1.54 | Deep Learning Feature 2 | 1.89 | Deep Learning Feature 1 | 1.30 | MIN RADIUS | 1.56 |
| Deep Learning Feature 46 | 1.49 | Deep Learning Feature 37 | 1.64 | LBP PERIPHERY 09 | 1.29 | LBP PERIPHERY 10 | 1.55 |
| Deep Learning Feature 48 | 1.48 | AREA | 1.62 | Deep Learning Feature 44 | 1.27 | Deep Learning Feature 59 | 1.49 |
| Deep Learning Feature 7 | 1.45 | LBP PERIPHERY 07 | 1.57 | Deep Learning Feature 63 | 1.19 | MEAN | 1.39 |

Feature importance represents the relative contribution of each morphometric or deep-learning feature to the corresponding AI-tumor cell morphotype.

Table 4: Definitions and interpretation of morphometric and image-derived features

| Feature | Feature category | Description | Interpretation for morphotype analysis |
|---|---|---|---|
| AREA | Size / shape | Area of the segmented cell or object. | Overall cell size. |
| PERIMETER | Size / shape | Length of the boundary of the segmented object. | Reflects cell size and contour complexity. |
| LONG AXIS | Size / shape | Length of the major (long) axis of the segmented object. | Reflects cell size and elongation. |
| SHORT AXIS | Size / shape | Length of the minor (short) axis of the segmented object. | Reflects cell width and shape. |
| MAX RADIUS | Size / shape | Maximum radial distance from the defined object center to its boundary. | Reflects cell size and contour irregularity. |
| MIN RADIUS | Size / shape | Minimum radial distance from the defined object center to its boundary. | Reflects cell shape and contour irregularity. |
| MAX FERET | Size / shape | Maximum Feret diameter: the largest caliper diameter of the segmented object. | Reflects the maximum dimension of the cell. |
| MIN FERET | Size / shape | Minimum Feret diameter: the smallest caliper diameter of the segmented object. | Reflects cell width and shape. |
| HU01 | Shape | First Hu invariant moment, a quantitative descriptor of object shape based on its spatial moments. | Captures aspects of cell shape while being relatively invariant to translation, scale, and rotation. |
| SMALL BLACK BLOB INTEGRAL | Texture / image pattern | Image-derived feature likely quantifying integrated signal associated with small dark regions or blobs within the cell image. | May capture intracellular dark structures or local image heterogeneity. |
| LBP PERIPHERY | Texture | Local Binary Pattern (LBP) feature/calculated from the peripheral region of the cell. | Captures a component of local texture at the cell periphery. |
| MEAN | Intensity | Mean pixel intensity within the segmented object. | Measures average image intensity of the cell. |
| STANDARD DEVIATION | Intensity / texture | Standard deviation of pixel intensity within the segmented object. | Measures heterogeneity of image intensity within the cell. |

LBP, local binary pattern. Hu invariant moments are quantitative shape descriptors derived from spatial moments and are relatively invariant to translation, scale, and rotation; n.a., not available.

Table 5: Oncogenes and tumor suppressor genes in regions with recurrent copy-number gains and losses

| Gene | Chr. | Cytoband | Gene type | Alteration | Patients, % | Patients, n |
|---|---|---|---|---|---|---|
| *MEF2D* | 1 | q22 | Oncogene | Gain | 50% | 5 |
| *NTRK1* | 1 | q23.1 | Oncogene | Gain | 50% | 5 |
| *PRCC* | 1 | q23.1 | Oncogene | Gain | 50% | 5 |
| *NCSTN* | 1 | q23.2 | Oncogene | Gain | 50% | 5 |
| *NUF2* | 1 | q23.3 | Oncogene | Gain | 50% | 5 |
| *COP1* | 1 | q25.1 | Oncogene | Gain | 50% | 5 |
| *PPARG* | 3 | p25.2 | Oncogene | Gain | 50% | 5 |
| *IRF2* | 4 | q35.1 | TSG | Loss | 60% | 6 |
| *ARHGAP26* | 5 | q31.3 | Oncogene | Gain | 50% | 5 |
| *FGF1* | 5 | q31.3 | Oncogene | Gain | 50% | 5 |
| *PDGFRB* | 5 | q32 | Oncogene | Gain | 50% | 5 |
| *CD74* | 5 | q33.1 | Oncogene | Gain | 50% | 5 |
| *GABRA6* | 5 | q34 | Oncogene | Gain | 50% | 5 |
| *DUSP4* | 8 | p12 | TSG | Loss | 50% | 5 |
| *ESCO2* | 8 | p21.1 | TSG | Loss | 60% | 6 |
| *NKX3-1* | 8 | p21.2 | TSG | Loss | 60% | 6 |
| *PPP2R2A* | 8 | p21.2 | TSG | Loss | 60% | 6 |
| *ATP6V1B2* | 8 | p21.3 | TSG | Loss | 60% | 6 |
| *LZTS1* | 8 | p21.3 | TSG | Loss | 60% | 6 |
| *MYC* | 8 | q24.21 | Oncogene | Gain | 60% | 6 |
| *CCND1* | 11 | q13.3 | Oncogene | Gain | 50% | 5 |
| *FGF19* | 11 | q13.3 | Oncogene | Gain | 50% | 5 |
| *FGF3* | 11 | q13.3 | Oncogene | Gain | 50% | 5 |
| *FGF4* | 11 | q13.3 | Oncogene | Gain | 50% | 5 |
| *EMSY* | 11 | q13.5 | Oncogene | Gain | 50% | 5 |
| *PAK1* | 11 | q13.5 | Oncogene | Gain | 50% | 5 |
| *ETS1* | 11 | q24.3 | Oncogene | Gain | 50% | 5 |
| *FLI1* | 11 | q24.3 | Oncogene | Gain | 50% | 5 |
| *BRCA2* | 13 | q13.1 | TSG | Loss | 50% | 5 |
| *PDS5B* | 13 | q13.1 | TSG | Loss | 50% | 5 |
| *DIS3* | 13 | q21.33 | TSG | Loss | 50% | 5 |
| *ERCC5* | 13 | q33.1 | TSG | Loss | 50% | 5 |
| *FGF14* | 13 | q33.1 | TSG | Loss | 50% | 5 |
| *PRKCB* | 16 | p12.2 | Oncogene | Gain | 50% | 5 |
| *PLCG2* | 16 | q23.3 | Oncogene | Gain | 50% | 5 |
| *FOXF1* | 16 | q24.1 | Oncogene | Gain | 50% | 5 |
| *MAP2K4* | 17 | p12 | TSG | Loss | 50% | 5 |

Frequency represents the proportion of patients with the indicated copy-number alteration; n.a., not available.

Table 6: Summary of variants observed across serial whole-exome sequencing samples

| Patient | No. timepoints | No. variants | Shared by all, n | Shared by $\geq$2 samples, n | Private variants, n | Shared by $\geq$2 samples, % |
|---|---|---|---|---|---|---|
| **ME002** | 6 | 22 | 0 | 4 | 18 | 18.2% |
| **ME003** | 2 | 15 | 1 | 1 | 14 | 6.7% |
| **ME006** | 4 | 43 | 0 | 5 | 38 | 11.6% |
| **ME007** | 3 | 8 | 0 | 0 | 8 | 0.0% |
| **ME008** | 2 | 31 | 16 | 16 | 15 | 51.6% |
| **ME014** | 3 | 19 | 7 | 8 | 11 | 42.1% |

Variants in OncoKB cnacer genes were detected by whole-exome sequencing of AI-tumor cell-enriched samples. Shared by all indicates variants detected at every available serial timepoint for a patient. Shared by $geq$2 samples indicates variants detected in at least two serial samples. Private variants were detected in only one serial sample.
